# Overexpression of HPRT1 is associated with poor prognosis in head and neck squamous cell carcinoma

**DOI:** 10.1101/2020.12.10.20246991

**Authors:** Mohsen Ahmadi, Pegah Mousavi, Negin Saffarzadeh, Fatemeh Hajiesmaeili, Leila Habibipour

## Abstract

Hypoxanthine phosphoribosyl transferase (*HPRT1*), as a salvage pathway enzyme, plays a crucial role in modulating the cell cycle and has been reported to be overexpressed in multiple cancers. Nevertheless, the relationship between the *HPRT1* and Head and Neck Squamous Cell Carcinomas (HNSCC) has not been investigated so far. We first evaluated the expression of *HPRT1* at transcriptomic and proteomic levels in tumor and healthy control tissues and its clinical value using The Cancer Genome Atlas (TCGA), Human Protein Atlas, Kaplan-Meier Plotter databases, GSE107591, and quantitative real-time PCR analysis. Then, we employed the COSMIC and cBioPortal databases to assess the mutations of the *HPRT1* gene and their association with survival outcomes of patients with HNSCC. Finally, we performed the functional enrichment analysis for *HPRT1* co-expressed genes in HNSCC utilizing the Enrichr database. The mRNA and protein expressions of *HPRT1* were significantly elevated in HNSCC compared with normal tissues. Besides, the upregulation of *HPRT1* expression was correlated with age, sex, pathological stage, and histological grades of HNSCC patients. Moreover, the increased expression of *HPRT1* in cancer tissues exhibited a strong capacity for being a promising biomarker for the diagnosis and prognosis of patients with HNSCC. The co-expressed genes of *HPRT1* were mainly enriched in several cancer-related processes such as DNA replication and cell cycle. The present study demonstrated that the overexpression of *HPRT1* is significantly correlated with the progression of HNSCC and may serve as a useful biomarker for the early detection and risk stratification of patients with HNSCC.

## Introduction

Head and Neck Squamous Cell Carcinoma (HNSCC) have been introduced as the sixth most common malignancy globally. HNSCC are originating from various subsites of the upper aerodigestive tract such as the oral cavity, larynx, and pharynx, paranasal sinuses, and nasal cavity [1-4]. Despite the current efforts for the risk stratification and treatment of HNSCC patients, the overall survival rate of these patients is unsatisfactory and has not significantly improved over the past decade may be due to late-stage diagnosis. The lack of effective means for clinical applications urgently demands the findings of more applicable biomarkers to improve the earlier detection and targeted therapies of patients with HNSCC [5-11].

Hypoxanthine phosphoribosyl transferase (*HPRT1*) transcripts the HPRT protein, a transferase enzyme that takes part in the cell cycle through modulation of guanine and inosine production in the salvage pathway [12, 13]. Although the *HPRT1* is broadly utilized as a housekeeping gene for many expression studies, growing evidence has ascertained the differential expression of *HPRT1* and its imperative role in quickly proliferating cells such as neoplasms due to elevated demand for nucleotides synthesis and consequently the *HPRT1* during the cell cycle [14, 15]. For instance, Townsend et al. have shown the overexpression of *HPRT1* in colorectal cancer samples when compared to healthy tissues [16]. Besides, others have demonstrated that the increased expression of *HPRT1* in endometrial cancer samples was correlated with patient’s survival outcomes [17]. Moreover, Sedano et al. also have discovered that upregulation of HPRT1 in tumor tissues could estimate the prognosis of patients with breast cancer [18]. These findings provoked us to appraise the *HPRT1* as a probable biomarker for HNSCC patients.

In the current study, we first analyzed the expression patterns of *HPRT1* in HNSCC using bioinformatics and laboratory investigations. We also assessed the diagnostic value and prognostic significance of *HPRT1* mRNA expression in patients with HNSCC. Besides, we conducted the mutation analysis for the *HPRT1* gene in HNSCC. Finally, we explored those genes that are closely associated with *HPRT1* in HNSCC and then performed the functional enrichment analysis for these co-expressed genes to provide a better understanding of *HPRT1* roles in HNSCC.

## Materials and Methods

### Expression analysis using the TCGA-HNSCC

The *HPRT1* mRNA expression and the data of several clinicopathological parameters of 520 HNSCC samples and 44 normal tissues in The Cancer Genome Atlas (TCGA) cohort were downloaded from data deposited in the University of California Santa Cruz (UCSC) Xena browser (https://xenabrowser.net/) [19]. The clinicopathological feature of patients of this cohort are shown in **Table S1**.

### Expression analysis using the GEO database

We compared the *HPRT1* mRNA expression in 23 paired HNSCC and non-cancerous tissues utilizing a previously published dataset (GSE107591) [20] obtained from the Gene Expression Omnibus (GEO) database [21]. *P*-value <0.05 was considered as statistically significant.

### Sample collection

We obtained a total of 45 tumor tissues and paired adjacent normal tissues from 45 patients with HNSCC undergoing routine surgical procedures in AmirAlam Hospital Complex in Tehran, Iran. It is worthy to note that none of the participants received treatment before the operation. The tissue samples were frozen immediately after surgery in liquid nitrogen and were stored at −80°C until RNA extraction. The demographic and clinical data of study participants are shown in **Table S2**. This study was approved by the Ethics Committee of Hormozgan University of Medical Sciences. Written informed consent was signed by all patients before involvement in this study.

### RNA isolation and qRT-PCR analysis

Total RNA from the patient’s tissue samples was isolated using TRIzolTM Reagent (Invitrogen, Carlsbad, CA, USA) based on the manufacturer’s protocol. The SuperScript IV Reverse Transcriptase cDNA Synthesis Kit (Roche, Germany) was employed for cDNA synthesis according to the manufacturer’s protocol. LightCycler 96 Real-Time PCR was used to the assessment of relative mRNA expression of *HPRT1* using SYBR Premix Ex Taq II (Tli RNaseH Plus) (TAKARA, Japan) under the manufacturer’s protocol. Each experiment was conducted in duplicate and the relative expression level was calculated using the -ddct method normalized with Succinate Dehydrogenase Complex Flavoprotein Subunit A (*SDHA)*. The primer sequences used in our study are deposited in **Table S3**.

### Immunohistochemistry staining

We assessed the expression of HPRT1 at protein levels in HNSCC and normal tissues using the Immunohistochemistry staining data collected from the online Human Protein Atlas (HPA) (https://www.proteinatlas.org/pathology) database [22-24]. We also analyzed the relationship between the HPRT1 protein expression and Overall Survival (OS) outcome in HNSCC patients. A log-rank *P*-value<0.05 was considered as statistically significant.

### Prognosis analysis

We obtained the information about the prognostic value of *HPRT1* mRNA expression in HNSCC via the Kaplan-Meier Plotter database [25]. A log-rank *P*-value<0.05 was considered as statistically significant.

### Mutation analysis

We introduced the Catalogue of Somatic Mutations in Cancer (COSMIC) database (http://cancer.sanger.ac.uk), to figure out the distribution and substitutions of the *HPRT1* gene mutations in HNSCC [26]. The data have been imported to Excel 2019 software to illustrate the Pie chart. Besides, we investigated the genetic alterations of the *HPRT1* gene and their impact on the survival outcomes of patients with HNSCC utilizing the data from the cBio Cancer Genomics Portal (http://cbioportal.org) database [27-29]. A log-rank *P*-value<0.05 was considered as statistically significant.

### Functional enrichment analysis

We investigated the co-expressed genes of *HPRT1* in HNSCC through the UALCAN database. genes with extremely low expression (median transcripts per million < 0.5) were removed, and only genes with a Pearson correlation coefficient ≥ 0.3 were involved for further analyses. We then applied the Enrichr (http://amp.pharm.mssm.edu/Enrichr) database to carry out the Gene Ontology (GO) and Kyoto Encyclopedia of Genes and Genomes (KEGG) pathway analysis [30]. The GO analysis contained Biological Processes (BP), Molecular Function (MF), and Cellular Component (CC). *P*-value<0.05 was defined as a cut-off criterion.

### Statistical Analysis

To compare the relative expression of *HPRT1* among different groups, a Student t-test or one-way analysis of variance (one-way ANOVA) tests were carried out as needed. Receiver operating characteristic (ROC) curves were illustrated to demonstrate the diagnostic utility of *HPRT1* mRNA expression for patients with HNSCC. The Area under the ROC Curve (AUC) closer to 1.0 reflected the test has the most diagnostic excellence. The maximum Youden index was employed as a cut-off point. *P*-value<0.05 was regarded as statistically significant. GraphPad Prism Software, Version 9.0.0, was utilized to perform statistical analysis.

### Database characteristics

The detailed information’s about each online database used for the present research has been described in the database characteristics section of the supplementary file.

## Results

### The *HPRT1* mRNA expression in HNSCC

The RNA-seq data revealed that the mRNA expression of HPRT1 was higher in HNSCC samples when compared to normal tissues (P-value<0.0001; Figure 1A).This was in accordance with our findings obtained from the GSE107591 dataset (P-value=0.022; Figure 1B). To confirm the results of our data mining, we assessed the HPRT1 mRNA expression levels in 45 paired HNSCC samples using Quantitative Real-Time Polymerase Chain Reaction (qRT-PCR) analysis. The results have concluded that the transcript levels of HPRT1 were apparently elevated in HNSCC than that in adjacent normal tissues (P-value<0.0001, Figure 1C).

**Figure 1.**
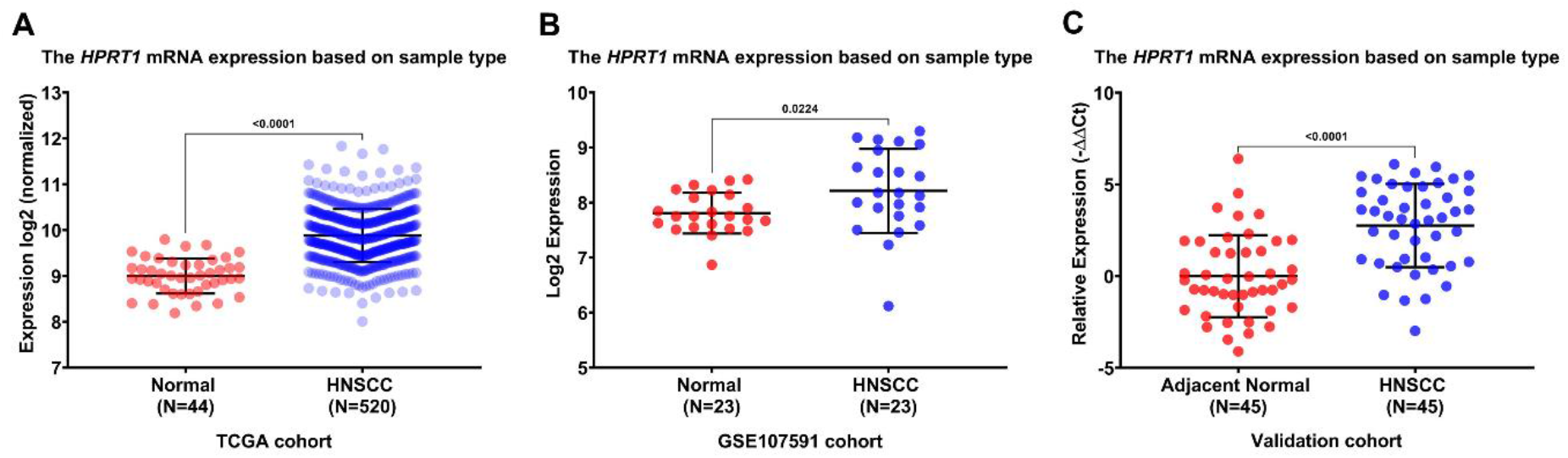
The expression analysis of *HPRT1* mRNA. The comparison of mRNA expression levels of *HPRT1* in 520 cancerous tissues and 44 healthy samples in the TCGA-HNSCC cohort(A). The *HPRT1* mRNA expression levels in 45 paired HNSCC and normal tissue specimens in the GSE25099 cohort utilizing the GEO database (B). The relative expression of *HPRT1* mRNA in 45 HNSCC samples and adjacent normal tissues in our validation cohort (C). All data are shown as the mean ± standard deviation. *P*-value<0.05 was regarded as statistically significant. TCGA: The Cancer Genome Atlas, HNSCC: Head and Neck Squamous Cell Carcinoma, GEO: Gene Expression Omnibus, N: Sample count.

### The relationship between the *HPRT1* mRNA expression and clinicopathological characteristics of patients with HNSCC

We further assessed the association of HPRT1 mRNA expression levels with clinicopathological features of patients in TCGA-HNSCC and our validation cohort. As presented in Figure 2A-H, the expression analysis determined that the overexpression of HPRT1 mRNA in cancer tissues was significantly correlated with the age, sex, pathological stage, and histological grade of participants of both studies. Table S2-3 represent the statistical significance of each comparison mentioned above for the TCGA-HNSCC and our validation cohorts, respectively.

**Figure 2.**
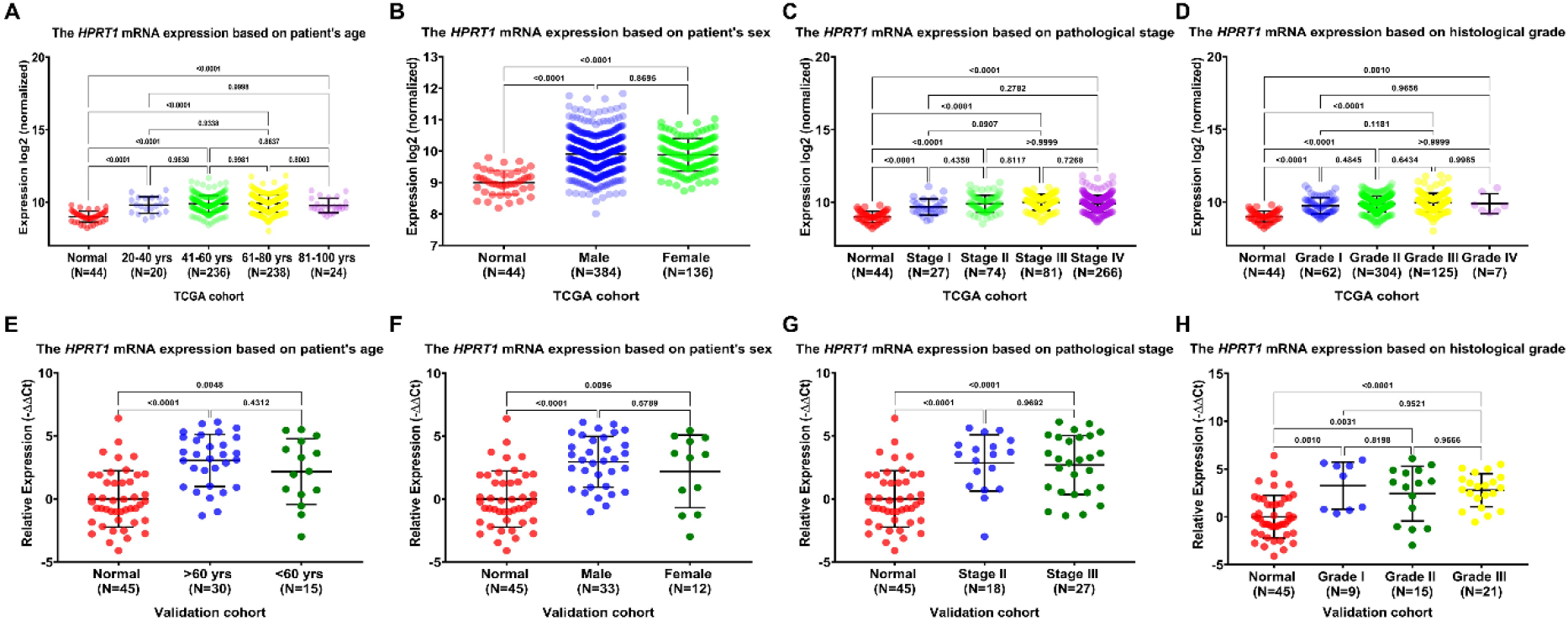
The association between the *HPRT1* mRNA expression levels and clinicopathological features. The correlation of *HPRT1* mRNA expression levels with patient’s age (A), gender (B), pathological stage (C), and histological grade (D) in the TCGA-HNSCC cohort. The relationship between the *HPRT1* mRNA expression levels and patient’s age (E), gender (F), pathological stage (G), and histological grade (H) in our validation cohort (qRT-PCR data). All data are shown as the mean ± standard deviation. *P*-value<0.05 was regarded as statistically significant. TCGA: The Cancer Genome Atlas, HNSCC: Head and Neck Squamous Cell Carcinoma, Qrt-PCR: Quantitative Real-Time Polymerase Chain Reaction, N: Sample count.

### The relationship between the HPRT1 protein expression and overall survival in HNSCC

We acquired the expression data of HPRT1 protein using the immunohistochemistry staining images from the HPA database. The findings showed the medium and high expression of HPRT1 protein in oral mucosa and HNSCC tissues, respectively (Figure 3A-B). We also used this database to discover the impact of HPRT1 protein expression on the survival time of HNSCC patients. We observed that the increased expression of HPRT1 protein was strongly correlated with poor OS in patients with HNSCC (P-value=0.000013; Figure 3C).

**Figure 3.**
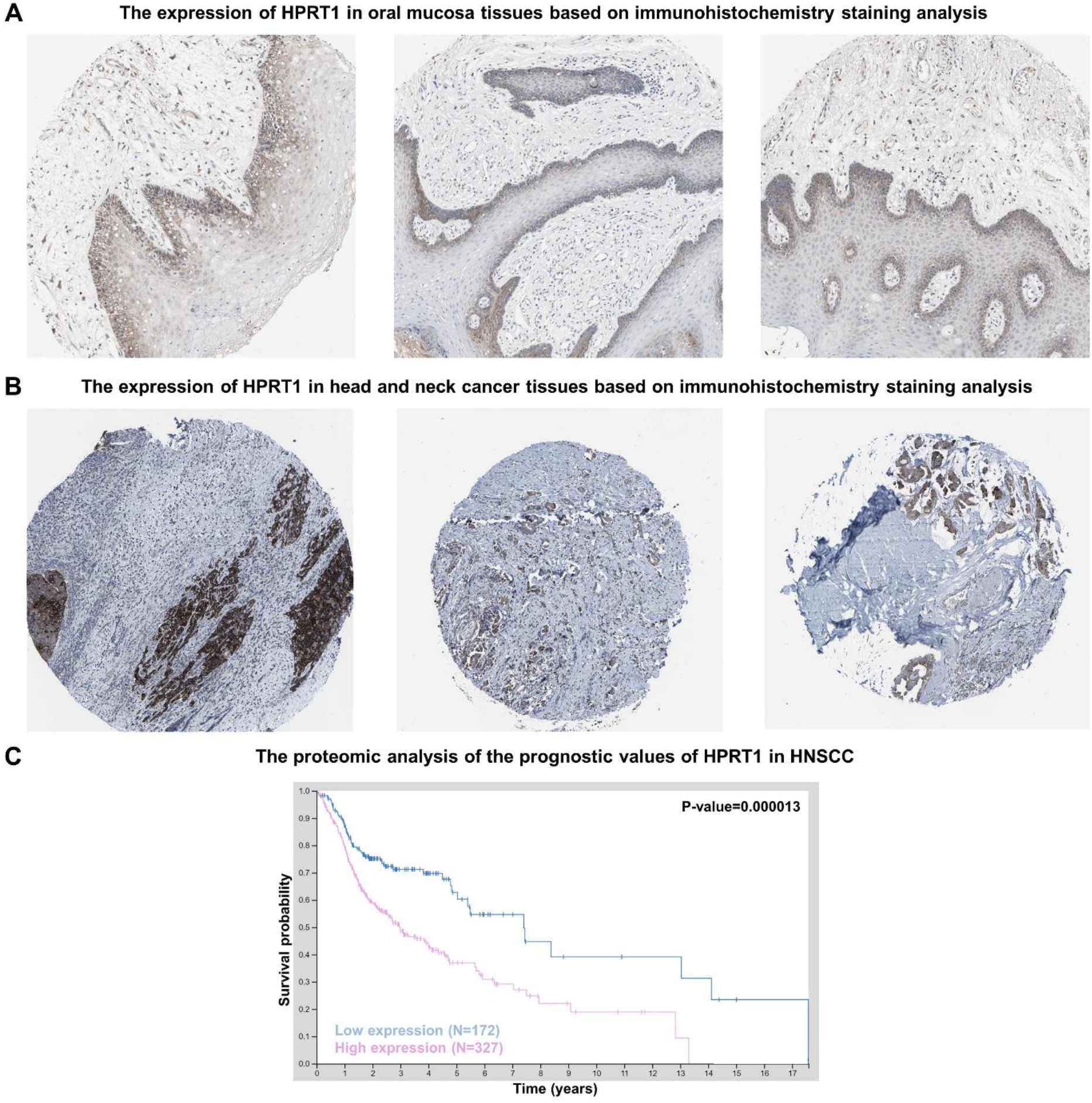
The proteomic analysis of HPRT1 protein expression in HNSCC using the data from the HPA database. The medium expression of HPRT1 protein in normal oral mucosa tissues (A). The high expression of HPRT1 protein in HNSCC tissues (B). The association of HPRT1 protein expression with survival outcome in patients with HNSCC (C). A log-rank *P*-value<0.05 was considered as statistically significant. HNSCC: Head and Neck Squamous Cell Carcinoma, HPA: Human Protein Atlas.

### The diagnostic utility of *HPRT1* mRNA expression for HNSCC

We appraised the diagnostic merit of HPRT1 mRNA expression for HNSCC through the calculation of the AUC in ROC curves. The findings demonstrated the diagnostic capacity of HPRT1 mRNA expression in HNSCC patients in TCGA cohort (AUC=0.901, P-value<0.0001, 95% CI=0.865-0.937; Figure 4A), GSE107591 cohort (AUC=0.701, P-value<0.019, 95% CI=0.542-0.859; Figure 4B), and in our validation cohort (AUC=0.806, P-value<0.0001, 95% CI=0.714– 0.897; Figure 4C).

**Figure 4.**
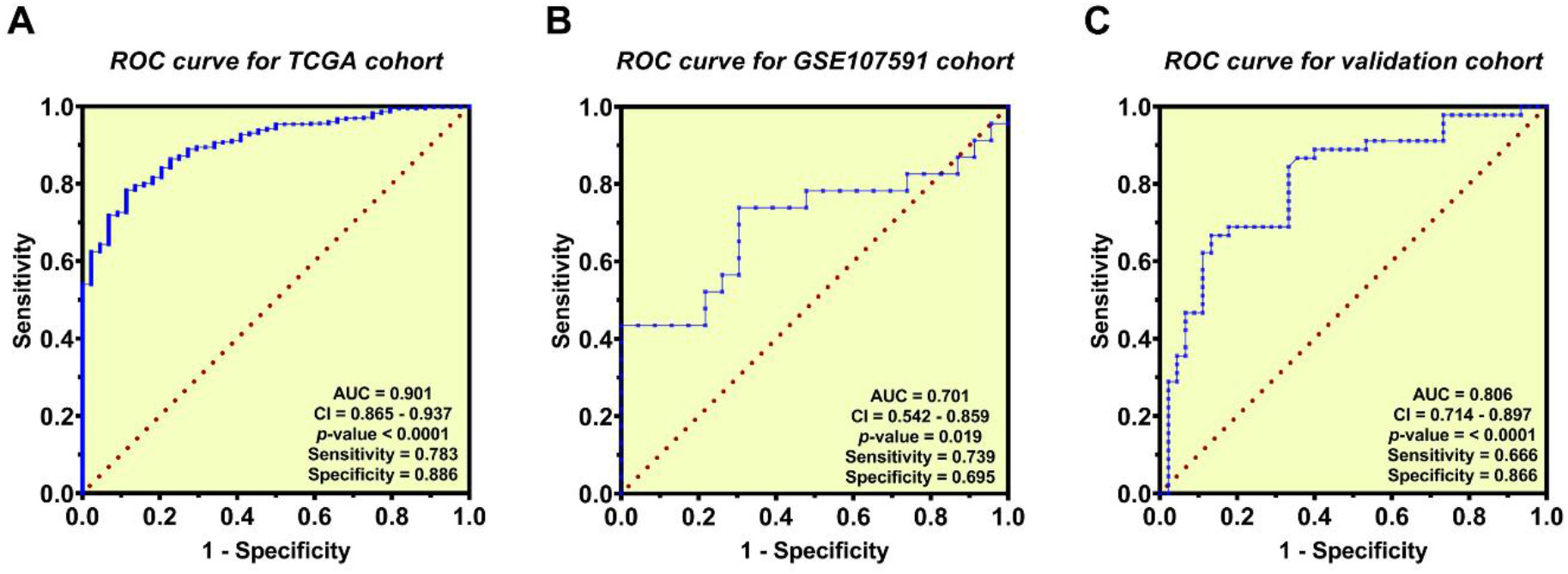
The diagnostic value of *HPRT1* mRNA expression in HNSCC using ROC curves analysis. ROC curves for the TCGA cohort (A). ROC curves for the GSE107591 cohort (B). ROC curves for the validation cohort (qRT-PCR data) (C). AUC: Area Under the ROC Curve, CI: Confidence Interval, ROC: Receiver Operating Characteristic, HNSCC: Head and Neck Squamous Cell Carcinoma, qRT-PCR: Quantitative Real-Time Polymerase Chain Reaction.

### Prognostic value of *HPRT1* mRNA expression for HNSCC

We evaluated the prognostic value of HPRT1 mRNA expression for HNSCC using the Kaplan-Meier Plotter database. The statistics suggested that higher levels of HPRT1 transcripts were correlated with the poor OS (P-value=1.3e-5; Figure 5A). However, there was no significant difference between Relapse Free Survival (RFS) in HNSCC patients with high expression levels of HPRT1 mRNA in comparison with those with low expression levels (P-value=0.22; Figure 5B).

**Figure 5.**
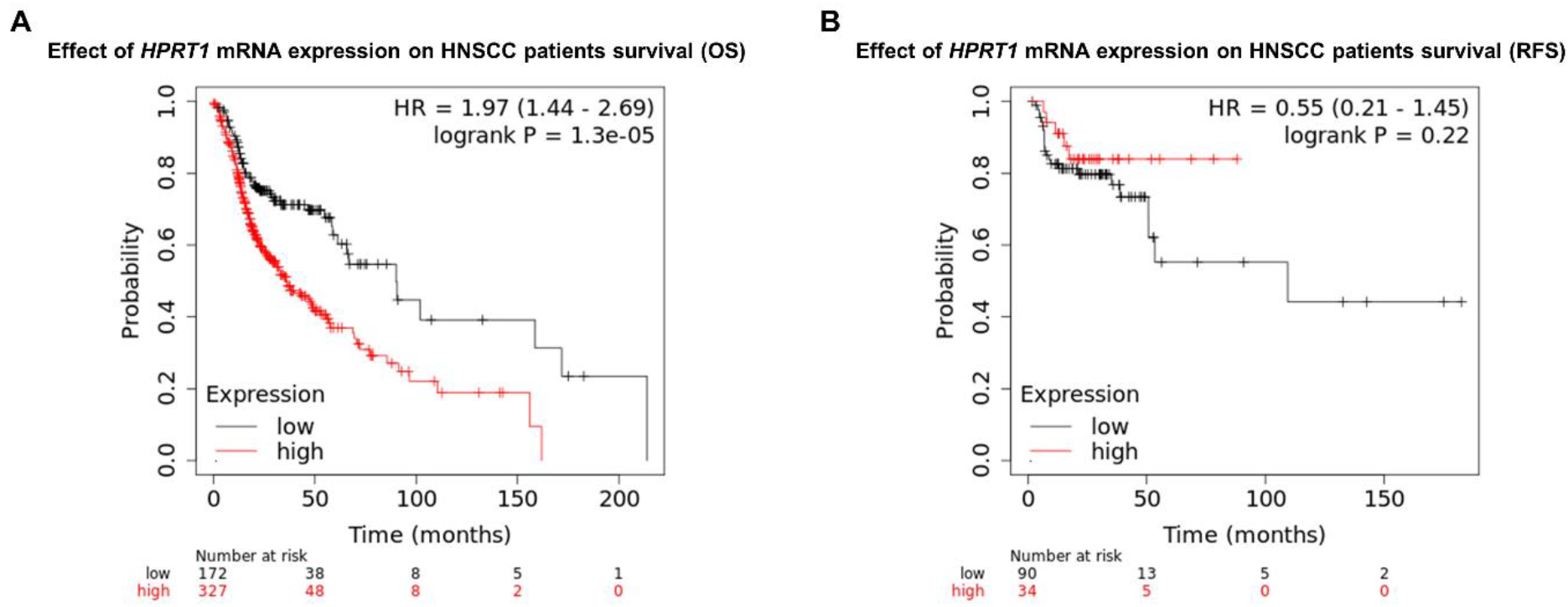
The prognostic significance of *HPRT1* mRNA expression in HNSCC using the data from the Kaplan-Meier Plotter database. The association between the *HPRT1* mRNA expression and OS in patients with HNSCC (A). The relationship between the *HPRT1* mRNA expression and RFS in HNSCC patients (B). A log-rank *P*-value<0.05 was considered as statistically significant. HNSCC: Head and Neck Squamous Cell Carcinoma, OS: Overall Survival, RFS: Relapse Free Survival.

### Mutation analysis of the *HPRT1* gene

The mutations of the HPRT1 gene in HNSCC samples were examined by the COSMIC database. The pie chart depicted that among four mutations in the HPRT1 gene, three were missense (75.00%) and one was synonymous (25.00%) substitutions (Figure 6A). There were 50.00% A>G, 25.00% G>A, and G>C mutations in the HPRT1 gene coding region (Figure 6A). We also explored the types of genetic alterations in the HPRT1 gene and their frequencies in 496 samples from the TCGA-HNSCC cohort using the cBioPortal database. As shown in Figure 6B, the HPRT1 gene was altered in 7 (1%) cases of HNSCC patients and the amplification was responsible for most changes. Moreover, using the “Survival” tab, we investigated the relationship between genetic alterations of the HPRT1 gene and survival times in patients with HNSCC. The Kaplan–Meier plot and log-rank test uncovered that genetic alterations of the HPRT1 gene were not associated with the Disease-Specific Survival (P-value=0.666; Figure S1A), Disease-Free Survival (P-value=0.194; Figure S1B), Progression-Free Survival (P-value=0.300; Figure S1C), and Overall Survival (P-value=0.130; Figure S1D) of HNSCC patients.

**Figure 6.**
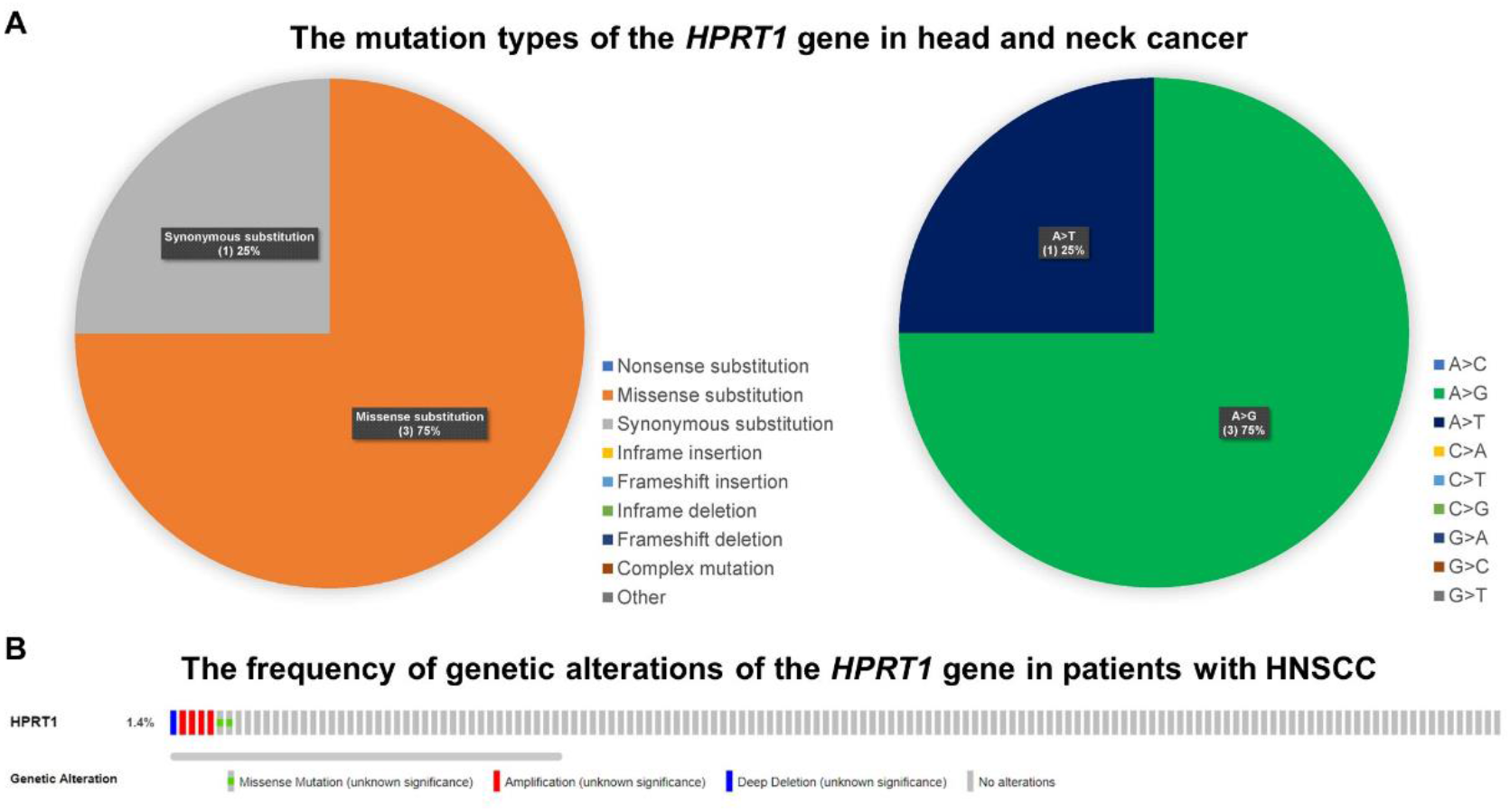
The mutation analysis for the *HPRT1* gene in HNSCC. The distribution and substitutions of mutations in the coding strand of the *HPRT1* gene in HNSCC based on the COSMIC database (A). The genetic alterations of the *HPRT1* gene and their frequencies in HNSCC patients according to the cBioPortal database (B). HNSCC: Head and Neck Squamous Cell Carcinoma, COSMIC: Catalogue of Somatic Mutations in Cancer.

### GO and KEGG pathway analysis for *HPRT1* co-expressed genes

To explore the potential roles of the HPRT1 in HNSCC, we first collected the co-expressed genes of the HPRT1 in HNSCC via the UALCAN database. Our data mining unveiled that 1933 genes were positively correlated with HPRT1 in HNSCC, as listed in Table S5. Additionally, we applied the Enrichr database to conduct the GO and KEGG pathway analysis for these co-expressed genes. The GO analysis unearthed that these genes were enriched in mRNA splicing, via spliceosome (GO:0000398), RNA splicing, via transesterification reactions with bulged adenosine as nucleophile (GO:0000377), mRNA processing (GO:0006397), DNA metabolic process (GO:0006259), RNA metabolic process (GO:0016070) in BP (Figure 7A), and RNA binding (GO:0003723), single-stranded DNA binding (GO:0003697), four-way junction DNA binding (GO:0000400), DNA secondary structure binding (GO:0000217), and DNA-dependent ATPase activity (GO:0008094) in MF (Figure 7B), and in the nucleolus (GO:0005730), condensed chromosome, centromeric region (GO:0000779), spliceosomal complex (GO:0005681), nuclear chromosome part (GO:0044454), and chromosomal region (GO:0098687) in CC (Figure 7C). The KEGG pathway analysis also indicated that these genes were enriched in RNA transport, Spliceosome, DNA replication, Cell cycle, and Ribosome biogenesis in eukaryotes pathways (Figure 7D).

**Figure 7.**
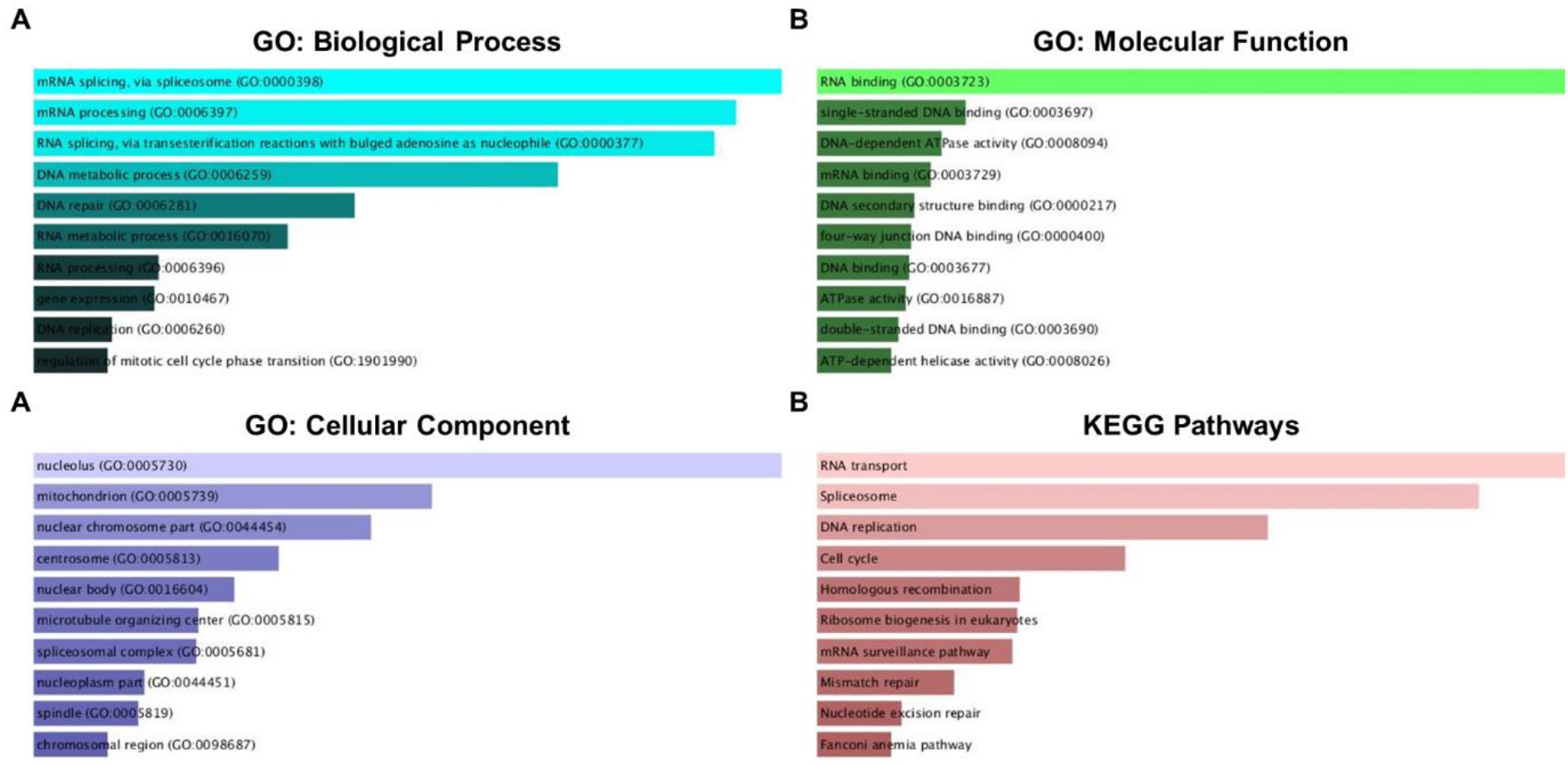
The data of GO and KEGG enrichment analysis. Based on the *HPRT1*-correlated genes, the enriched information of Biological Process (A), Molecular Function (B), Cellular Component and (C) in GO analysis and KEGG pathways (D) were obtained using the Enrichr database. GO: Gene Ontology, KEGG: Kyoto Encyclopedia of Genes and Genomes.

## Discussion

The *HPRT1* aberrant expression has been documented in several cancers [15]. But the relationship between the *HPRT1* and HNSCC is unknown. To our knowledge, our report is the first to investigate the expression of the *HPRT1* at transcriptomic and proteomic levels as well as its genetic alterations, diagnostic and prognostic merit, and probable functions in the head and neck squamous cancer. We hope that the findings of the present research advance the current knowledge, enhance therapeutic targets, and strengthen the accuracy of diagnosis and prognosis for HNSCC patients.

In this study, we initially employed the data from TCGA-HNSCC cohort and GSE107591 dataset to demonstrate the *HPRT1* mRNA expression in HNSCC. The results showed that the *HPRT1* mRNA expression was significantly upregulated in cancer tissues comparing to healthy control samples, which was consistent with the findings of our qRT-PCR analysis for 45 paired HNSCC and normal tissues. We also assessed the association of the *HPRT1* mRNA levels with the sex, age, pathological stage, and histological grades of HNSCC patients in TCGA-HNSCC and our validation cohorts. The gender-based and individuals’ age-based differences have been contributed to the clinical presentations and prognosis of patients with HNSCC, respectively [31-34]. The sex-based analysis for both cohorts indicated the significant overexpression of *HPRT1* gene level in tumor samples of both genders compared with the normal tissues. However, there were no statistically significant differences between transcript levels of the *HPRT1* gene in cancerous tissues of males and females. Besides, we observed that there was no significant association between the *HPRT1* mRNA expression levels and the patients’ age in tumor tissues in both cohorts. These may reflect that the patients’ gender and age probably did not influence the expression levels of *HPTR1* mRNA in cancer tissues. We also discovered that the *HPRT1* mRNA expression levels were remarkably elevated in advanced pathological stages and histological grades of HNSCC tissues in comparison with healthy controls in both cohorts. These observations suggested that the *HPRT1* may take part in the pathogenesis of head and neck squamous cancer. According to the small size of tissue specimens in our validation cohort, we were unable to evaluate the expression of the HPRT1 in protein levels. Therefore, the HPA was introduced, and the IHC staining images have revealed the medium and high expression levels of the HPRT1 protein in the normal oral mucosa and head and neck cancer tissues, respectively. All together, these findings unearthed that expression of HPRT1 was significantly increased in HNSCC tissues.

One of the most difficult challenges about HNSCC patients is the time and procedure of screening and diagnosis. It has been declared that the delayed diagnosis and treatment of patients with HNSCC aggravate the prognosis outcomes and increase the undesired morbidity and mortality of cancer patients. Hence, findings of new biomarkers would boost the clinical outcomes of patients with HNSCC [35-38]. In the current research, we carried out the ROC curves analysis to appraise the diagnostic utility of the *HPRT1* mRNA expression levels for HNSCC. The data from TCGA (AUC=0.901), GSE107591 (AUC=0.701), and our validation cohort (AUC=0.806) concluded that expression levels of the *HPRT1* gene could be considered as a useful marker for diagnosis of HNSCC patients. Moreover, the prognosis data obtained from the Kaplan-Meier Plotter and HPA databases disclosed that the higher expression levels of the HPRT1 in cancer tissues were significantly correlated with inferior survival time of patients with HNSCC, in line with the findings of previous studies that suggested the dysregulation of the *HPRT1* expression was is associated with poor survival outcomes of breast and endometrial cancer [17, 18]. In sum, these results may indicate the great value of the *HPRT1* tissue expression for diagnosis and risk stratification of patients with HNSCC.

It is widely accepted that the accumulation of genetic alterations plays a causal role in the tumorigenesis of HNSCC [39]. The further assessments using the COSMIC and cBioPortal databases unveiled that genetic alterations of the *HPRT1* gene was rare in HNSCC patients (7/496), and the missense mutations were responsible for the most type of genetic aberrations in the *HPRT1* gene coding strand. Besides, the genetic alterations of the *HPRT1* gene were not associated with survival outcomes of patients with HNSCC. Furthermore, we uncovered that the *HPRT1* and its co-expressed genes were enriched in the RNA transport, Spliceosome, DNA replication, Cell cycle, and Ribosome biogenesis in eukaryotes pathways. We hypothesized that the *HPRT1* mediates the progression of HNSCC through these pathways. Noteworthy, others have proposed that the *HPRT1*, as a crucial component of the purine salvage pathway, plays a role in the mediation of the proliferation, autophagy, and apoptosis-related processes in cancer cells [40-42]. Nevertheless, the exact function of the *HPRT1* in these pathways is ambiguous and additional studies are required to clarify the underlying mechanisms of the *HPRT1* in these pathways.

## Conclusion

Overall, the findings of the current research study indicated that overexpression of the *HPRT1* is significantly correlated with the progression of HNSCC. Besides, the observed upregulation of the *HPRT1* expression in tumor tissues may be a valuable indicator for diagnosis and prognosis biomarker for patients with HNSCC. However, further investigations and clinical trials are mandatory to elucidate the involvement of the *HPRT1* in HNSCC.

## Supporting information

The clinicopathological feature of patients of this cohort are shown in Table S1.

## Data Availability

The data of TCGA were obtained from the website of university of California Santa Cruz (UCSC) Xena browser (https ://xenab rowser.net/). The data of the GSE107591 dataset were downloaded from the website of the GEO database (https://www.ncbi.nlm.nih.gov/geo/). The data of the validation cohort used for this research are available from the corresponding author upon request.

## Authors’ contributions

NS collected the patient’s samples and laboratory materials. MA and FH performed the laboratory experiments. MA conducted the data extraction and statistical analysis and initially drafted the paper. PM contributed to the design of the study, supervised all investigations. NS, PM, FH, and LH edited the manuscript. MA and PM checked the final version of the manuscript. All authors have contributed to, read, and approved the final manuscript for submission.

## Acknowledgments

This study was supported by Hormozgan University of Medical Sciences and Booali Medical Diagnostic Laboratory.

## Data Availability Statement

The data of TCGA were obtained from the website of niversity of California Santa Cruz (UCSC) Xena browser (https://xenabrowser.net/). The data of GSE107591 dataset were downloaded from the website of the GEO database (https://www.ncbi.nlm.nih.gov/geo/). The data of the validation cohort used for this research are available from the corresponding author upon request.

## Abbreviations

AUC: Area Under the ROC Curve
BP: Biological Processes
CC: Cellular Component
COSMIC: Catalogue of Somatic Mutations in Cancer
GEO: Gene Expression Omnibus
GO: Gene Ontology
HNSCC: Head and Neck Squamous Cell Carcinomas
HPA: Human Protein Atlas
HPRT1: Hypoxanthine phosphoribosyl transferase
KEGG: Kyoto Encyclopedia of Genes and Genomes
MF: Molecular Function
OS: Overall Survival
qRT-PCR: Quantitative Real-Time Polymerase Chain Reaction
RFS: Relapse Free Survival
ROC: Receiver Operating Characteristic
SDHA: Succinate Dehydrogenase Complex Flavoprotein Subunit A
TCGA: The Cancer Genome Atlas
UCSC: University of California Santa Cruz

## References

[1] S. Jia, M. Zhang, Y. Li, L. Zhang, W. Dai, MAGE-A11 Expression Predicts Patient Prognosis in Head and Neck Squamous Cell Carcinoma, Cancer Manag Res 12 (2020) 1427–1435.

[2] J.C.-H. Hsieh, H.-M. Wang, M.-H. Wu, K.-P. Chang, P.-H. Chang, C.-T. Liao, C.-T. Liau, Review of emerging biomarkers in head and neck squamous cell carcinoma in the era of immunotherapy and targeted therapy, Head & Neck 41(S1) (2019) 19–45.

[3] P. Economopoulou, R. de Bree, I. Kotsantis, A. Psyrri, Diagnostic Tumor Markers in Head and Neck Squamous Cell Carcinoma (HNSCC) in the Clinical Setting, Frontiers in Oncology 9(827) (2019).

[4] E. Alsahafi, K. Begg, I. Amelio, N. Raulf, P. Lucarelli, T. Sauter, M. Tavassoli, Clinical update on head and neck cancer: molecular biology and ongoing challenges, Cell Death & Disease 10(8) (2019) 540.

[5] G. Fan, Y. Tu, N. Wu, H. Xiao, The expression profiles and prognostic values of HSPs family members in Head and neck cancer, Cancer Cell International 20(1) (2020) 220.

[6] Y. Jin, Y. Yang, Bioinformatics-based discovery of PYGM and TNNC2 as potential biomarkers of head and neck squamous cell carcinoma, Bioscience Reports 39(7) (2019).

[7] A. Jemal, F. Bray, M.M. Center, J. Ferlay, E. Ward, D. Forman, Global cancer statistics, CA Cancer J Clin 61(2) (2011) 69–90.

[8] F. Bray, J. Ferlay, I. Soerjomataram, R.L. Siegel, L.A. Torre, A. Jemal, Global cancer statistics 2018: GLOBOCAN estimates of incidence and mortality worldwide for 36 cancers in 185 countries, CA Cancer J Clin 68(6) (2018) 394–424.

[9] P. Jiang, Y. Zhang, J.A.S H. Wang, Adoptive cell transfer after chemotherapy enhances survival in patients with resectable HNSCC, Int Immunopharmacol 28(1) (2015) 208–14.

[10] K. Payne, J. Brooks, R. Spruce, N. Batis, G. Taylor, P. Nankivell, H. Mehanna, Circulating Tumour Cell Biomarkers in Head and Neck Cancer: Current Progress and Future Prospects, Cancers (Basel) 11(8) (2019) 1115.

[11] Y.-t. Chen, J.-n. Yao, Y.-t. Qin, K. Hu, F. Wu, Y.-y. Fang, Biological role and clinical value of miR-99a-5p in head and neck squamous cell carcinoma (HNSCC): A bioinformatics-based study, FEBS Open Bio 8(8) (2018) 1280–1298.

[12] R.J. Monnat Jr, T.A. Chiaverotti, A.F. Hackmann, G.A. Maresh, Molecular structure and genetic stability of human hypoxanthine phosphoribosyltransferase (HPRT) gene duplications, Genomics 13(3) (1992) 788–796.

[13] R.J. Torres, J.G. Puig, Hypoxanthine-guanine phosophoribosyltransferase (HPRT) deficiency: Lesch-Nyhan syndrome, Orphanet journal of rare diseases 2(1) (2007) 48.

[14] M.H. Townsend, A.M. Felsted, Z.E. Ence, S.R. Piccolo, R.A. Robison, K. O’Neill, Elevated expression of hypoxanthine guanine phosphoribosyltransferase within malignant tissue, Cancer Clin Oncol 6(6) (2017) 19–34.

[15] M.H. Passey, A.M. Felsted, Z.E. Ence, S.R. Piccolo, K.L. Neill, R.A. Robison, Abstract 2536: Unique HPRT1 upregulation in malignant tissue: Potential use as diagnostic biomarker, Cancer Research 78(13 Supplement) (2018) 2536.

[16] M.H. Townsend, A.M. Felsted, W. Burrup, R.A. Robison, K.L. O’Neill, Examination of Hypoxanthine Guanine Phosphoribosyltransferase as a biomarker for colorectal cancer patients, Mol Cell Oncol 5(4) (2018) e1481810.

[17] M.H. Townsend, Z.E. Ence, A.M. Felsted, A.C. Parker, S.R. Piccolo, R.A. Robison, K.L. O’Neill, Potential new biomarkers for endometrial cancer, Cancer Cell International 19(1) (2019) 19.

[18] M. J Sedano, E. I Ramos, R. Choudhari, A. L Harrison, R. Subramani, R. Lakshmanaswamy, M. Zilaie, S.S. Gadad, Hypoxanthine Phosphoribosyl Transferase 1 Is Upregulated, Predicts Clinical Outcome and Controls Gene Expression in Breast Cancer, Cancers (Basel) 12(6) (2020) 1522.

[19] M. Goldman, B. Craft, J. Zhu, D. Haussler, The UCSC Xena system for cancer genomics data visualization and interpretation, AACR, 2017.

[20] L. Verduci, M. Ferraiuolo, A. Sacconi, F. Ganci, J. Vitale, T. Colombo, P. Paci, S. Strano, G. Macino, N. Rajewsky, G. Blandino, The oncogenic role of circPVT1 in head and neck squamous cell carcinoma is mediated through the mutant p53/YAP/TEAD transcription-competent complex, Genome Biol 18(1) (2017) 237.

[21] T. Barrett, D.B. Troup, S.E. Wilhite, P. Ledoux, D. Rudnev, C. Evangelista, I.F. Kim, A. Soboleva, M. Tomashevsky, R. Edgar, NCBI GEO: mining tens of millions of expression profiles—database and tools update, Nucleic acids research 35(Suppl_1) (2007) D760–D765.

[22] M. Uhlén, L. Fagerberg, B.M. Hallström, C. Lindskog, P. Oksvold, A. Mardinoglu, Å. Sivertsson, C. Kampf, E. Sjöstedt, A. Asplund, Tissue-based map of the human proteome, Science 347(6220) (2015).

[23] P.J. Thul, L. Åkesson, M. Wiking, D. Mahdessian, A. Geladaki, H.A. Blal, T. Alm, A. Asplund, L. Björk, L.M. Breckels, A subcellular map of the human proteome, Science 356(6340) (2017).

[24] M. Uhlen, C. Zhang, S. Lee, E. Sjöstedt, L. Fagerberg, G. Bidkhori, R. Benfeitas, M. Arif, Z. Liu, F. Edfors, A pathology atlas of the human cancer transcriptome, Science 357(6352) (2017).

[25] Á. Nagy, A. Lánczky, O. Menyhárt, B. Győrffy, Validation of miRNA prognostic power in hepatocellular carcinoma using expression data of independent datasets, Scientific reports 8(1) (2018) 1–9.

[26] S.A. Forbes, D. Beare, H. Boutselakis, S. Bamford, N. Bindal, J. Tate, C.G. Cole, S. Ward, E. Dawson, L. Ponting, COSMIC: somatic cancer genetics at high-resolution, Nucleic acids research 45(D1) (2017) D777–D783.

[27] P. Wu, Z.J. Heins, J.T. Muller, L. Katsnelson, I. de Bruijn, A.A. Abeshouse, N. Schultz, D. Fenyö, J. Gao, Integration and analysis of CPTAC proteomics data in the context of cancer genomics in the cBioPortal, Molecular & Cellular Proteomics 18(9) (2019) 1893–1898.

[28] J. Gao, B.A. Aksoy, U. Dogrusoz, G. Dresdner, B. Gross, S.O. Sumer, Y. Sun, A. Jacobsen, R. Sinha, E. Larsson, Integrative analysis of complex cancer genomics and clinical profiles using the cBioPortal, Science signaling 6(269) (2013) pl1–pl1.

[29] E. Cerami, J. Gao, U. Dogrusoz, B.E. Gross, S.O. Sumer, B.A. Aksoy, A. Jacobsen, C.J. Byrne, M.L. Heuer, E. Larsson, The cBio cancer genomics portal: an open platform for exploring multidimensional cancer genomics data, AACR, 2012.

[30] M.V. Kuleshov, M.R. Jones, A.D. Rouillard, N.F. Fernandez, Q. Duan, Z. Wang, S. Koplev, S.L. Jenkins, K.M. Jagodnik, A. Lachmann, Enrichr: a comprehensive gene set enrichment analysis web server 2016 update, Nucleic acids research 44(W1) (2016) W90–W97.

[31] T. Hoang, C. Zhang, H.M. Geye, T. Speer, M. Yu, P.A. Wiederholt, J.C. Kakuske, A.M. Traynor, J.F. Cleary, S.H. Dailey, Gender associations in head and neck squamous cell carcinoma (HNSCC), American Society of Clinical Oncology, 2012.

[32] S.T. Massa, L.M. Cass, S. Challapalli, Z. Zahirsha, M. Simpson, G. Ward, N. Osazuwa-Peters, Demographic predictors of head and neck cancer survival differ in the elderly, The Laryngoscope 129(1) (2019) 146–153.

[33] F. Qadir, A. Lalli, H.H. Dar, S. Hwang, H. Aldehlawi, H. Ma, H. Dai, A. Waseem, M.-T. Teh, Clinical correlation of opposing molecular signatures in head and neck squamous cell carcinoma, BMC cancer 19(1) (2019) 830.

[34] R. Hollows, W. Wei, J.B. Cazier, H. Mehanna, G. Parry, G. Halford, P. Murray, Association between loss of Y chromosome and poor prognosis in male head and neck squamous cell carcinoma, Head & Neck 41(4) (2019) 993–1006.

[35] H.W. Schutte, F. Heutink, D.J. Wellenstein, G.B. van den Broek, F.J.A. van den Hoogen, H.A.M. Marres, C.M.L. van Herpen, J. Kaanders, T. Merkx, R.P. Takes, Impact of Time to Diagnosis and Treatment in Head and Neck Cancer: A Systematic Review, Otolaryngol Head Neck Surg 162(4) (2020) 446–457.

[36] A. Coca-Pelaz, R.P. Takes, K. Hutcheson, N.F. Saba, M. Haigentz, C.R. Bradford, R. de Bree, P. Strojan, V.J. Lund, W.M. Mendenhall, I.J. Nixon, M. Quer, A. Rinaldo, A. Ferlito, Head and Neck Cancer: A Review of the Impact of Treatment Delay on Outcome, Advances in Therapy 35(2) (2018) 153–160.

[37] Q. Li, Z. Shen, Z. Wu, Y. Shen, H. Deng, C. Zhou, H. Liu, High P4HA1 expression is an independent prognostic factor for poor overall survival and recurrent-free survival in head and neck squamous cell carcinoma, Journal of Clinical Laboratory Analysis 34(3) (2020) e23107.

[38] V. Budach, I. Tinhofer, Novel prognostic clinical factors and biomarkers for outcome prediction in head and neck cancer: a systematic review, The Lancet Oncology 20(6) (2019) e313–e326.

[39] S.V. Puram, I. Tirosh, A.S. Parikh, A.P. Patel, K. Yizhak, S. Gillespie, C. Rodman, C.L. Luo, E.A. Mroz, K.S. Emerick, Single-cell transcriptomic analysis of primary and metastatic tumor ecosystems in head and neck cancer, Cell 171(7) (2017) 1611-1624. e24.

[40] M. Camici, M. Garcia-Gil, R. Pesi, S. Allegrini, M.G. Tozzi, Purine-metabolising enzymes and apoptosis in cancer, Cancers (Basel) 11(9) (2019) 1354.

[41] J. Yin, W. Ren, X. Huang, J. Deng, T. Li, Y. Yin, Potential Mechanisms Connecting Purine Metabolism and Cancer Therapy, Frontiers in Immunology 9(1697) (2018).

[42] E. Villa, E.S. Ali, U. Sahu, I. Ben-Sahra, Cancer Cells Tune the Signaling Pathways to Empower de Novo Synthesis of Nucleotides, Cancers (Basel) 11(5) (2019) 688.

