## Supplementary material for "Overexpression of HPRT1 is associated with poor prognosis in head and neck squamous cell carcinoma": The clinicopathological feature of patients of this cohort are shown in Table S1.

### Database characteristics:

The Gene Expression Omnibus (GEO) database (https://www.ncbi.nlm.nih.gov/geo/) is an useful resource of available gene expression data that can be combined and investigated to derive new theories and knowledge [1].

University of California Santa Cruz (UCSC) Xena browser (https ://xenab rowser.net/) is a useful resource for analyzing and visualization of multi-omic data and associated clinical and phenotypic annotations [2].

The Human Protein Atlas (HPA) (https://www.proteinatlas.org/pathology) is the largest and most comprehensive database for proteins in tissues and cells, providing an invaluable data for exploration of expression profile at a single-cell resolution [3].

The Catalogue of Somatic Mutations in Cancer (COSMIC) database (http://cancer.sanger.ac.uk), is a high-resolution web tool for exploring the impact of somatic mutations in all kind of human cancers [4].

The cBio Cancer Genomics Portal (http://cbioportal.org), is an open-access resource for cancer genomic datasets [5].

The Kaplan–Meier plotter (http://kmplot.com/analysis), is an online platform that can be utilized to assess the influence of 54,675 genes on survival outcomes of patients in 21 cancer types [6, 7].

Enrichr (http://amp.pharm.mssm.edu/Enrichr) database is a comprehensive platform for curated gene sets and a search engine that provides biological knowledge for further biological hypothesis [8].

### Tables:

**Table S1.** Clinicopathological feature of patients in the TCGA-HNSCC cohort.

| Clinicopathological parameter | Category | N | Percent |
|---|---|---|---|
| Tissue specimens | HNSCC | 520 | 92 |
| | Non-cancerous | 44 | 8 |
| Age | 20-40 | 24 | 4 |
| | 41-60 | 249 | 44 |
| | 61-80 | 262 | 47 |
| | 81-100 | 27 | 5 |
| Sex | Female | 150 | 27 |
| | Male | 414 | 73 |
| Stage | I | 27 | 5 |
| | II | 74 | 14 |
| | III | 81 | 16 |
| | IV | 266 | 51 |
| | NA | 71 | 14 |
| Grade | G1 | 62 | 12 |
| | G2 | 304 | 59 |
| | G3 | 125 | 24 |
| | G4 | 7 | 1 |
| | NA | 18 | 4 |

TCGA: The Cancer Genome Atlas, HNSCC: Head and Neck Squamous Cell Carcinoma, NA: Not Available, N: Sample count.

**Table S2.** Clinicopathological feature of patients in the validation cohort.

| Clinicopathological parameter | Category | M | Percent |
|---|---|---|---|
| Tissue specimens | HNSCC | 45 | 50 |
| | Non-cancerous | 45 | 50 |
| Age | <60 | 30 | 68 |
| | ≥60 | 15 | 32 |
| Sex | Female | 33 | 73 |
| | Male | 12 | 27 |
| Stage | I-II | 18 | 40 |
| | III-IV | 27 | 40 |
| Grade | G1 | 9 | 20 |
| | G2 | 15 | 33 |
| | G3 | 21 | 47 |

TCGA: The Cancer Genome Atlas, HNSCC: Head and Neck Squamous Cell Carcinoma, NA: Not Available, N: Sample count.

**Table S3.** Primer sequences used in this study.

| qRT-PCR primers | Type of primers | Sequences |
|---|---|---|

| | | |
|---|---|---|
| HPRT1 (target gene) | Forward primer | 5'-TGGCGTCGTGATTAGTGATG-3' |
| HPRT1 (target gene) | Reverse primer | 5'-ACAGAGGGCTACAATGTGATG-3' |
| SDHA (reference gene) | Forward primer | 5'-CTTGCCAGGACCTAGAGTTTGT-3' |
| SDHA (reference gene) | Reverse primer | 5'-CTCTCCACGACATCCTTCCG-3' |

**Table S4.** The association between *HPRT1* mRNA expression and clinicopathological features of HNSCC patients in TCGA cohort.

| **Comparison** | **N** | | **Mean Difference** | ***P*-values** | **Significant** |
|---|---|---|---|---|---|
| | **A1** | **A2** | | | |
| HNSCC vs. Normal | 520 | 44 | 0.8821 | <0.0001 | yes |
| Male vs. Normal | 384 | 45 | 0.9098 | <0.0001 | yes |
| Female vs. Normal | 136 | 45 | 0.8821 | <0.0001 | yes |
| Female vs. Male | 136 | 384 | -0.02764 | 0.8695 | ns |
| Age (20-40 yrs) vs. Normal | 20 | 44 | 0.7990 | <0.0001 | yes |
| Age (41-60 yrs) vs. Normal | 236 | 44 | 0.8870 | <0.0001 | yes |
| Age (61-80 yrs) vs. Normal | 238 | 44 | 0.9028 | <0.0001 | yes |
| Age (81-100 yrs) vs. Normal | 24 | 44 | 0.7683 | <0.0001 | yes |
| Age (41-60 yrs) vs. Age (20-40 yrs) | 236 | 20 | 0.08800 | 0.9630 | ns |
| Age (61-80 yrs) vs. Age (20-40 yrs) | 238 | 20 | 0.1038 | 0.9338 | ns |
| Age (81-100 yrs) vs. Age (20-40 yrs) | 24 | 20 | -0.03076 | 0.9998 | ns |
| Age (61-80 yrs) vs. Age (41-60 yrs) | 238 | 236 | 0.01576 | 0.9981 | ns |
| Age (81-100 yrs) vs. Age (41-60 yrs) | 24 | 236 | -0.1188 | 0.8637 | ns |
| Age (81-100 yrs) vs. Age (61-80 yrs) | 24 | 238 | -0.1345 | 0.8003 | ns |
| Stage I vs. Normal | 27 | 44 | 0.6792 | <0.0001 | yes |
| Stage II vs. Normal | 74 | 44 | 0.8926 | <0.0001 | yes |
| Stage III vs. Normal | 81 | 44 | 0.9906 | <0.0001 | yes |
| Stage IV vs. Normal | 266 | 44 | 0.9025 | <0.0001 | yes |
| Stage II vs. Stage I | 74 | 27 | 0.2134 | 0.4358 | ns |
| Stage III vs. Stage I | 81 | 27 | 0.3114 | 0.0907 | ns |
| Stage IV vs. Stage I | 266 | 27 | 0.2233 | 0.2782 | ns |
| Stage III vs. Stage II | 81 | 74 | 0.09797 | 0.8117 | ns |
| Stage IV vs. Stage II | 266 | 74 | 0.009883 | >0.9999 | ns |
| Stage IV vs. Stage III | 266 | 81 | -0.08808 | 0.7268 | ns |
| Grade I vs. Normal | 62 | 44 | 0.7456 | <0.0001 | yes |
| Grade II vs. Normal | 304 | 44 | 0.8725 | <0.0001 | yes |
| Grade III vs. Normal | 125 | 44 | 0.9546 | <0.0001 | yes |
| Grade IV vs. Normal | 7 | 44 | 0.8923 | 0.0010 | yes |
| Grade II vs. Grade I | 304 | 62 | 0.1269 | 0.4845 | ns |
| Grade III vs. Grade I | 125 | 62 | 0.2090 | 0.1181 | ns |
| Grade IV vs. Grade I | 21 | 62 | 0.1467 | 0.9656 | ns |
| Grade III vs. Grade II | 125 | 304 | 0.08211 | 0.6434 | ns |
| Grade IV vs. Grade II | 7 | 304 | 0.01985 | >0.9999 | ns |
| Grade IV vs. Grade III | 7 | 125 | -0.06226 | 0.9985 | ns |

TCGA: The Cancer Genome Atlas, HNSCC: Head and Neck Squamous Cell Carcinoma, N: Sample count, A1: Attribute 1, A1: Attribute 2, ns: not significant.

**Table S5.** The association between *HPRT1* mRNA expression and clinicopathological features of HNSCC patients in our validation cohort.

| Comparison | N | | Mean Difference | *P*-values | Significant |
|---|---|---|---|---|---|
| | A1 | A2 | | | |
| HNSCC vs. Normal | 45 | 45 | 2.758 | <0.0001 | yes |
| Male vs. Normal | 33 | 45 | 2.961 | <0.0001 | yes |
| Female vs. Normal | 12 | 45 | 2.201 | 0.0096 | yes |
| Female vs. Male | 12 | 33 | -0.7597 | 0.5789 | ns |
| Age (>60 yrs) vs. Normal | 30 | 45 | 3.053 | <0.0001 | yes |
| Age (<60 yrs) vs. Normal | 15 | 45 | 2.169 | 0.0048 | yes |
| Age (<60 yrs) vs. Age (>60 yrs) | 15 | 30 | -0.8834 | 0.4312 | ns |
| Stage II vs. Normal | 18 | 45 | 2.857 | <0.0001 | yes |
| Stage III vs. Normal | 27 | 45 | 2.693 | <0.0001 | yes |
| Stage III vs. Stage II | 27 | 18 | -0.1643 | 0.9692 | ns |
| Grade I vs. Normal | 9 | 45 | 3.259 | 0.001 | yes |
| Grade II vs. Normal | 15 | 45 | 2.425 | 0.0031 | yes |
| Grade III vs. Normal | 21 | 45 | 2.782 | <0.0001 | yes |
| Grade II vs. Grade I | 15 | 9 | -0.8336 | 0.8198 | ns |
| Grade III vs. Grade I | 21 | 9 | -0.4774 | 0.9521 | ns |
| Grade III vs. Grade II | 21 | 15 | 0.3562 | 0.9666 | ns |

HNSCC: Head and Neck Squamous Cell Carcinoma, N: Sample count, A1: Attribute 1, A1: Attribute 2, ns: not significant.

**Table S6.** The co-expressed genes of *HPRT1* in HNSCC, collected from the UALCAN database.

| Genes | Pearson CC |
|---|---|
| VBP1 | 0.66 |
| PGRMC1 | 0.63 |
| RBMX2 | 0.62 |
| DKC1 | 0.62 |
| PSMD10 | 0.59 |
| VMA21 | 0.59 |
| NKRF | 0.59 |
| PRPS1 | 0.57 |
| CHAC2 | 0.56 |
| ZNF280C | 0.56 |
| UTP14A | 0.56 |
| FAM122B | 0.56 |
| ALG13 | 0.56 |
| GLA | 0.55 |
| SLC25A5 | 0.55 |
| NKAP | 0.55 |
| AIFM1 | 0.54 |
| VRK1 | 0.54 |
| PTGES3 | 0.54 |
| NONO | 0.53 |

| | |
|---|---|
| XPO1 | 0.53 |
| PHF5A | 0.53 |
| TIMM8A | 0.53 |
| CCDC59 | 0.53 |
| DSCC1 | 0.52 |
| HTATSF1 | 0.52 |
| CHRNA5 | 0.52 |
| SLC25A17 | 0.52 |
| MORF4L2 | 0.52 |
| TCEAL4 | 0.52 |
| MST4 | 0.52 |
| KIF4A | 0.51 |
| CDKN3 | 0.51 |
| C15orf23 | 0.51 |
| ORC6L | 0.51 |
| UPF3B | 0.51 |
| MMGT1 | 0.51 |
| HNRNPC | 0.51 |
| EMD | 0.51 |
| SFRS7 | 0.51 |
| RBMX | 0.51 |
| GPN3 | 0.51 |
| FAM127B | 0.51 |
| GLRX5 | 0.5 |
| UBE2A | 0.5 |
| XRCC6 | 0.5 |
| SSB | 0.5 |
| OCRL | 0.5 |
| MAP7D3 | 0.5 |
| MRPL30 | 0.5 |
| PHF6 | 0.5 |
| ADSL | 0.5 |
| CXorf56 | 0.49 |
| CENPI | 0.49 |
| EPR1 | 0.49 |
| SLC25A14 | 0.49 |
| MTA1 | 0.49 |
| RAD54B | 0.49 |
| CEBPZ | 0.49 |
| KLHL13 | 0.48 |
| CCNB2 | 0.48 |
| WDR67 | 0.48 |
| DPY30 | 0.48 |
| ERH | 0.48 |
| MNAT1 | 0.48 |
| BUB1 | 0.48 |

| | |
|---|---|
| ERCC6L | 0.48 |
| NUP37 | 0.48 |
| UBE2T | 0.48 |
| SFRS9 | 0.48 |
| MTHFD2 | 0.48 |
| MAD2L1 | 0.48 |
| RAD51L3 | 0.48 |
| ST13 | 0.48 |
| MCTS1 | 0.48 |
| HSPD1 | 0.48 |
| CBX3 | 0.48 |
| BOLA3 | 0.48 |
| MTCP1NB | 0.48 |
| DHX9 | 0.48 |
| PGK1 | 0.48 |
| SFRS3 | 0.48 |
| MAPKAPK5 | 0.48 |
| LAS1L | 0.48 |
| C14orf143 | 0.47 |
| NUF2 | 0.47 |
| CNIH4 | 0.47 |
| ILF2 | 0.47 |
| SPC25 | 0.47 |
| CCT4 | 0.47 |
| PAICS | 0.47 |
| CDK1 | 0.47 |
| C12orf48 | 0.47 |
| UBA2 | 0.47 |
| PNO1 | 0.47 |
| ARMCX5 | 0.47 |
| FANCB | 0.47 |
| SIP1 | 0.47 |
| DLGAP5 | 0.47 |
| ACTL6A | 0.47 |
| THOC2 | 0.47 |
| STAG2 | 0.46 |
| C1GALT1C1 | 0.46 |
| CALM2 | 0.46 |
| TMEM97 | 0.46 |
| FAM72A | 0.46 |
| KIF23 | 0.46 |
| ZC3H8 | 0.46 |
| PPAT | 0.46 |
| CACYBP | 0.46 |
| ENOX2 | 0.46 |
| UTP18 | 0.46 |

| | |
|---|---|
| CETN2 | 0.46 |
| FAM10A4 | 0.46 |
| EXO1 | 0.46 |
| ATPBD4 | 0.46 |
| CENPA | 0.46 |
| HNRNPA3P1 | 0.46 |
| HSP90AA1 | 0.46 |
| ABCB7 | 0.46 |
| ATP11C | 0.46 |
| PPM1G | 0.46 |
| KIF18A | 0.46 |
| BUB1B | 0.46 |
| C16orf61 | 0.46 |
| TDP1 | 0.45 |
| POLR2D | 0.45 |
| CNIH | 0.45 |
| LOC728024 | 0.45 |
| TRMT61B | 0.45 |
| KIF4B | 0.45 |
| HAT1 | 0.45 |
| MRPL42 | 0.45 |
| C17orf75 | 0.45 |
| TROAP | 0.45 |
| RAN | 0.45 |
| NCAPH | 0.45 |
| CENPN | 0.45 |
| HNRNPA2B1 | 0.45 |
| TCEA1 | 0.45 |
| DAP3 | 0.45 |
| CSTF2 | 0.45 |
| CECR5 | 0.45 |
| ZCRB1 | 0.45 |
| DDX24 | 0.45 |
| C2orf3 | 0.45 |
| TPRKB | 0.45 |
| ZWILCH | 0.45 |
| CDC45 | 0.44 |
| FAM58A | 0.44 |
| SUV39H2 | 0.44 |
| FAM54A | 0.44 |
| PSMA3 | 0.44 |
| GABPB1 | 0.44 |
| TAF1A | 0.44 |
| BCCIP | 0.44 |
| NHP2L1 | 0.44 |
| APEX1 | 0.44 |

| | |
|---|---|
| FAM72B | 0.44 |
| UBE2N | 0.44 |
| FAM96A | 0.44 |
| ACP1 | 0.44 |
| HIBADH | 0.44 |
| G2E3 | 0.44 |
| CDK4 | 0.44 |
| GTSE1 | 0.44 |
| RRP15 | 0.44 |
| TSEN15 | 0.44 |
| UBL4A | 0.44 |
| WDHD1 | 0.44 |
| NCRNA00183 | 0.44 |
| HNRNPH2 | 0.44 |
| MCM10 | 0.44 |
| IMMT | 0.44 |
| C1orf131 | 0.44 |
| ECT2 | 0.44 |
| TFAM | 0.44 |
| DDX50 | 0.43 |
| PPHLN1 | 0.43 |
| MRPL35 | 0.43 |
| KPNA2 | 0.43 |
| PPIL5 | 0.43 |
| CHD1L | 0.43 |
| SFXN1 | 0.43 |
| PCBP2 | 0.43 |
| RNASEH1 | 0.43 |
| NUP62CL | 0.43 |
| YY1 | 0.43 |
| XRCC3 | 0.43 |
| SPATS2 | 0.43 |
| DDX55 | 0.43 |
| TTK | 0.43 |
| KIF14 | 0.43 |
| SNW1 | 0.43 |
| CXorf40B | 0.43 |
| PYCR2 | 0.43 |
| TMLHE | 0.43 |
| DBF4 | 0.43 |
| C12orf24 | 0.43 |
| PARP2 | 0.43 |
| AHSA1 | 0.43 |
| C2orf56 | 0.43 |
| ACTR6 | 0.43 |
| SRP9 | 0.43 |

| | |
|---|---|
| FOXM1 | 0.43 |
| NEK2 | 0.43 |
| MECP2 | 0.43 |
| RPAP3 | 0.43 |
| PRDX3 | 0.43 |
| MRPS23 | 0.43 |
| DARS | 0.43 |
| HMGB3 | 0.43 |
| PBK | 0.43 |
| PDCD10 | 0.43 |
| HNRNPA3 | 0.43 |
| PWP1 | 0.43 |
| USP39 | 0.43 |
| RFC5 | 0.43 |
| PTPLAD1 | 0.43 |
| TATDN1 | 0.43 |
| HNRNPL | 0.43 |
| AURKA | 0.43 |
| HNRPLL | 0.43 |
| RAP2C | 0.43 |
| PPP1CC | 0.43 |
| MED30 | 0.43 |
| CUL4B | 0.42 |
| TOMM22 | 0.42 |
| ZC3H14 | 0.42 |
| CDCA7 | 0.42 |
| PDZD11 | 0.42 |
| UCK2 | 0.42 |
| SMNDC1 | 0.42 |
| WDR43 | 0.42 |
| MKI67IP | 0.42 |
| SUPT16H | 0.42 |
| FMR1 | 0.42 |
| C14orf104 | 0.42 |
| CHEK2 | 0.42 |
| POLG2 | 0.42 |
| MRPL3 | 0.42 |
| PRKRA | 0.42 |
| LRRC37B2 | 0.42 |
| ARL6IP6 | 0.42 |
| MTERFD1 | 0.42 |
| NAP1L1 | 0.42 |
| CEP55 | 0.42 |
| HAX1 | 0.42 |
| ARHGAP11A | 0.42 |
| RFC2 | 0.42 |

| | |
|---|---|
| THUMPD2 | 0.42 |
| HAUS2 | 0.42 |
| HNRNPK | 0.42 |
| CCNA2 | 0.42 |
| RNF113A | 0.42 |
| COX7A2L | 0.42 |
| XRCC6BP1 | 0.42 |
| PRIM2 | 0.42 |
| PPP3R1 | 0.42 |
| HSPE1 | 0.42 |
| NLE1 | 0.42 |
| BLM | 0.42 |
| DENR | 0.42 |
| RAD51 | 0.42 |
| FAM72D | 0.42 |
| NFYB | 0.42 |
| HNRNPU | 0.42 |
| DEPDC1B | 0.42 |
| NEIL3 | 0.42 |
| AGK | 0.42 |
| OSGIN2 | 0.42 |
| DBF4B | 0.42 |
| PNN | 0.42 |
| PAPOLA | 0.42 |
| ANP32E | 0.42 |
| MSH6 | 0.42 |
| BOD1 | 0.42 |
| SNRPG | 0.42 |
| GGCT | 0.42 |
| R3HDM1 | 0.42 |
| TUBA1B | 0.42 |
| IARS2 | 0.42 |
| GMPS | 0.42 |
| CENPL | 0.41 |
| GNG10 | 0.41 |
| CSE1L | 0.41 |
| SFRS2 | 0.41 |
| NCBP2 | 0.41 |
| CCT6P1 | 0.41 |
| TMX1 | 0.41 |
| AMZ2 | 0.41 |
| HNRNPR | 0.41 |
| HMGN4 | 0.41 |
| SEC11A | 0.41 |
| C16orf80 | 0.41 |
| NOP58 | 0.41 |

| | |
|---|---|
| TRMT2B | 0.41 |
| SNAPC5 | 0.41 |
| RAE1 | 0.41 |
| MAGT1 | 0.41 |
| LRPPRC | 0.41 |
| CHCHD3 | 0.41 |
| SUMO2 | 0.41 |
| LOC649330 | 0.41 |
| CRIPT | 0.41 |
| IKBKG | 0.41 |
| METTL2A | 0.41 |
| MTIF2 | 0.41 |
| KIAA1467 | 0.41 |
| HJURP | 0.41 |
| PNPT1 | 0.41 |
| FANCM | 0.41 |
| DEPDC4 | 0.41 |
| C14orf109 | 0.41 |
| KIF18B | 0.41 |
| TRA2B | 0.41 |
| AURKAPS1 | 0.41 |
| TIPIN | 0.41 |
| DLEU2 | 0.41 |
| MRPL9 | 0.41 |
| C12orf32 | 0.41 |
| CDCA5 | 0.41 |
| SF3B14 | 0.41 |
| TOMM20 | 0.41 |
| MRPS35 | 0.41 |
| C14orf169 | 0.41 |
| MORC4 | 0.41 |
| UBR7 | 0.41 |
| SET | 0.41 |
| ANP32A | 0.41 |
| SPAG5 | 0.41 |
| DARS2 | 0.41 |
| HOXA10 | 0.41 |
| TRIP13 | 0.41 |
| MGAT2 | 0.41 |
| SLC9A6 | 0.41 |
| TOP2A | 0.41 |
| FANCI | 0.41 |
| RLIM | 0.41 |
| OLA1 | 0.41 |
| TCEAL1 | 0.41 |
| RHEB | 0.41 |

| | |
|---|---|
| CDC6 | 0.41 |
| WDR12 | 0.41 |
| NOL11 | 0.41 |
| NUCKS1 | 0.41 |
| NAE1 | 0.41 |
| UTP6 | 0.41 |
| KIAA0101 | 0.41 |
| PSMD14 | 0.41 |
| MTHFS | 0.41 |
| UIMC1 | 0.4 |
| FAM199X | 0.4 |
| C12orf11 | 0.4 |
| LOC493754 | 0.4 |
| CCT3 | 0.4 |
| ASF1A | 0.4 |
| SUMO1P3 | 0.4 |
| GDI1 | 0.4 |
| RFC4 | 0.4 |
| C17orf42 | 0.4 |
| KRR1 | 0.4 |
| ANP32B | 0.4 |
| RACGAP1 | 0.4 |
| LSM12 | 0.4 |
| IGBP1 | 0.4 |
| RSL24D1 | 0.4 |
| CCDC138 | 0.4 |
| PARP1 | 0.4 |
| CDCA3 | 0.4 |
| PTCD3 | 0.4 |
| TIAL1 | 0.4 |
| PRC1 | 0.4 |
| UTP23 | 0.4 |
| LOC723972 | 0.4 |
| C1orf27 | 0.4 |
| SSRP1 | 0.4 |
| BRCC3 | 0.4 |
| CRCP | 0.4 |
| DNA2 | 0.4 |
| PTDSS1 | 0.4 |
| ZNF26 | 0.4 |
| MTFMT | 0.4 |
| ZDHHC9 | 0.4 |
| TBPL1 | 0.4 |
| CCDC41 | 0.4 |
| CDCA4 | 0.4 |
| CKS2 | 0.4 |

| | |
|---|---|
| XRCC2 | 0.4 |
| CDC123 | 0.4 |
| E2F6 | 0.4 |
| DPY19L4 | 0.4 |
| NUDCD1 | 0.4 |
| COPS3 | 0.4 |
| BZW2 | 0.4 |
| NOL10 | 0.4 |
| COQ3 | 0.4 |
| ASPM | 0.4 |
| ZBTB33 | 0.4 |
| ARL6IP1 | 0.4 |
| NF1 | 0.4 |
| SLC35B1 | 0.4 |
| C1orf135 | 0.4 |
| DEPDC1 | 0.4 |
| DIABLO | 0.4 |
| METAP2 | 0.4 |
| YDJC | 0.4 |
| H2AFZ | 0.4 |
| COX4NB | 0.4 |
| KPNB1 | 0.4 |
| OTUD6B | 0.4 |
| FAM189B | 0.4 |
| TIMM17A | 0.4 |
| C10orf119 | 0.4 |
| VKORC1L1 | 0.4 |
| C3orf26 | 0.4 |
| RAD51AP1 | 0.4 |
| HNRNPA1L2 | 0.4 |
| FLVCR1 | 0.4 |
| PLK1 | 0.4 |
| SMC4 | 0.4 |
| CDCA7L | 0.4 |
| MTBP | 0.4 |
| IAH1 | 0.4 |
| BRIX1 | 0.39 |
| EPT1 | 0.39 |
| F8A1 | 0.39 |
| C1QBP | 0.39 |
| SEPHS1 | 0.39 |
| TBCCD1 | 0.39 |
| CCNB1IP1 | 0.39 |
| RBBP7 | 0.39 |
| TXNRD1 | 0.39 |
| BRMS1L | 0.39 |

| | |
|---|---|
| KIAA1586 | 0.39 |
| TERF1 | 0.39 |
| VAMP7 | 0.39 |
| SFPQ | 0.39 |
| NUP93 | 0.39 |
| CCDC104 | 0.39 |
| HDGFRP3 | 0.39 |
| WBSCR22 | 0.39 |
| MTHFD1 | 0.39 |
| NGFRAP1 | 0.39 |
| TBCE | 0.39 |
| LAMP2 | 0.39 |
| NUP205 | 0.39 |
| RBM28 | 0.39 |
| MATR3 | 0.39 |
| SNUPN | 0.39 |
| AIMP1 | 0.39 |
| DNAJC2 | 0.39 |
| HDAC2 | 0.39 |
| MSH2 | 0.39 |
| DTL | 0.39 |
| COX11 | 0.39 |
| GMNN | 0.39 |
| KLHL7 | 0.39 |
| C16orf63 | 0.39 |
| ZFAND1 | 0.39 |
| FKBP3 | 0.39 |
| CIRH1A | 0.39 |
| HMGB1 | 0.39 |
| WDR75 | 0.39 |
| SFRS13A | 0.39 |
| DRG1 | 0.39 |
| NLN | 0.39 |
| HELLS | 0.39 |
| KHDRBS1 | 0.39 |
| ATAD5 | 0.39 |
| CDCA8 | 0.39 |
| H2AFV | 0.39 |
| PRPSAP2 | 0.39 |
| C11orf84 | 0.39 |
| GINS2 | 0.39 |
| ASB3 | 0.39 |
| HAUS8 | 0.39 |
| TFB2M | 0.39 |
| ZNF449 | 0.39 |
| KIF15 | 0.39 |

| | |
|---|---|
| IRAK1 | 0.39 |
| DDX18 | 0.39 |
| NUP133 | 0.39 |
| PRELID2 | 0.39 |
| DCAF13 | 0.39 |
| LIN9 | 0.39 |
| STAMBP | 0.39 |
| TMEM206 | 0.39 |
| TMEM60 | 0.39 |
| MRPL19 | 0.39 |
| C3orf33 | 0.39 |
| NOB1 | 0.39 |
| FAM64A | 0.39 |
| SFRS1 | 0.39 |
| TSPAN13 | 0.39 |
| EXOSC2 | 0.39 |
| NGDN | 0.39 |
| AURKB | 0.39 |
| CBX1 | 0.39 |
| BUB3 | 0.39 |
| HSPA14 | 0.39 |
| RNASEH2B | 0.39 |
| WDYHV1 | 0.39 |
| RSL1D1 | 0.39 |
| CENPO | 0.39 |
| SPG21 | 0.39 |
| ORC2L | 0.39 |
| RBM17 | 0.39 |
| TOR3A | 0.39 |
| PSMD11 | 0.39 |
| TSN | 0.39 |
| MAP4K3 | 0.39 |
| C15orf44 | 0.39 |
| TAZ | 0.39 |
| CDK2 | 0.39 |
| ADSS | 0.39 |
| USP10 | 0.39 |
| PUS1 | 0.39 |
| FEN1 | 0.39 |
| C1orf31 | 0.39 |
| GSG2 | 0.39 |
| CPSF6 | 0.39 |
| SUPV3L1 | 0.39 |
| PCGF6 | 0.39 |
| TMEM164 | 0.39 |
| POLR2H | 0.39 |

| | |
|---|---|
| IPO9 | 0.38 |
| PRMT5 | 0.38 |
| RFT1 | 0.38 |
| SLC6A8 | 0.38 |
| MPHOSPH9 | 0.38 |
| MTCP1 | 0.38 |
| RCN2 | 0.38 |
| STARD7 | 0.38 |
| LBR | 0.38 |
| FXR1 | 0.38 |
| ZMYM3 | 0.38 |
| LSM5 | 0.38 |
| SLC4A1AP | 0.38 |
| BAG2 | 0.38 |
| RSRC2 | 0.38 |
| ATP2B1 | 0.38 |
| WASF1 | 0.38 |
| G3BP1 | 0.38 |
| TXNDC12 | 0.38 |
| HNRNPD | 0.38 |
| RBM34 | 0.38 |
| NUPL2 | 0.38 |
| HDAC8 | 0.38 |
| TMEM14B | 0.38 |
| TATDN3 | 0.38 |
| NOM1 | 0.38 |
| GPI | 0.38 |
| FAM122C | 0.38 |
| WDR33 | 0.38 |
| CEP290 | 0.38 |
| RBM41 | 0.38 |
| MYBBP1A | 0.38 |
| ISCA2 | 0.38 |
| DIAPH2 | 0.38 |
| SNRPF | 0.38 |
| SPR | 0.38 |
| RRS1 | 0.38 |
| NOC3L | 0.38 |
| CCNK | 0.38 |
| BCAS2 | 0.38 |
| RPA3 | 0.38 |
| C14orf106 | 0.38 |
| DDX47 | 0.38 |
| MRPL47 | 0.38 |
| NSL1 | 0.38 |
| OBFC2B | 0.38 |

| | |
|---|---|
| SUV39H1 | 0.38 |
| SNAP47 | 0.38 |
| SNRNP27 | 0.38 |
| BRCA1 | 0.38 |
| SCHIP1 | 0.38 |
| AHCTF1 | 0.38 |
| FH | 0.38 |
| NLK | 0.38 |
| NDUFB5 | 0.38 |
| TAF1 | 0.38 |
| DUT | 0.38 |
| KPNA4 | 0.38 |
| YAF2 | 0.38 |
| NUDT21 | 0.38 |
| YWHAE | 0.38 |
| MRPL17 | 0.38 |
| C6orf115 | 0.38 |
| RAD21 | 0.38 |
| UBXN2A | 0.38 |
| ATXN10 | 0.38 |
| CHRAC1 | 0.38 |
| PDXP | 0.38 |
| TFB1M | 0.38 |
| CHML | 0.38 |
| UBE2CBP | 0.38 |
| NIPSNAP1 | 0.38 |
| GTF2H4 | 0.38 |
| GPN1 | 0.38 |
| SELT | 0.38 |
| NPM1 | 0.38 |
| LOC100130932 | 0.38 |
| DUSP12 | 0.38 |
| TFAP4 | 0.38 |
| ZNF639 | 0.38 |
| COPS2 | 0.38 |
| HNRNPH3 | 0.38 |
| POLE2 | 0.38 |
| EIF2AK1 | 0.38 |
| HMMR | 0.38 |
| KIAA1804 | 0.38 |
| SGOL2 | 0.38 |
| C14orf101 | 0.38 |
| TCP1 | 0.37 |
| EIF3J | 0.37 |
| NCL | 0.37 |
| LAPTM4B | 0.37 |

| | |
|---|---|
| GPHN | 0.37 |
| TTC5 | 0.37 |
| FBXO5 | 0.37 |
| FUBP1 | 0.37 |
| SRRT | 0.37 |
| U2AF2 | 0.37 |
| XPNPEP3 | 0.37 |
| ZC3H15 | 0.37 |
| SMARCB1 | 0.37 |
| CKS1B | 0.37 |
| STRAP | 0.37 |
| TP53BP1 | 0.37 |
| FAM92A1 | 0.37 |
| C8orf76 | 0.37 |
| CEP152 | 0.37 |
| PA2G4P4 | 0.37 |
| MAGOHB | 0.37 |
| OIP5 | 0.37 |
| TYW1 | 0.37 |
| MDH1 | 0.37 |
| KIF11 | 0.37 |
| FAM133B | 0.37 |
| CCDC99 | 0.37 |
| ZBTB2 | 0.37 |
| MND1 | 0.37 |
| PPIL4 | 0.37 |
| TMEM170A | 0.37 |
| FANCA | 0.37 |
| RANBP1 | 0.37 |
| EPCAM | 0.37 |
| SLC38A6 | 0.37 |
| RBM45 | 0.37 |
| TMED8 | 0.37 |
| MCM7 | 0.37 |
| TAF15 | 0.37 |
| NDUFA6 | 0.37 |
| KIF20B | 0.37 |
| EEF1E1 | 0.37 |
| NUP54 | 0.37 |
| DLEU1 | 0.37 |
| METTL5 | 0.37 |
| PSRC1 | 0.37 |
| DNAJB11 | 0.37 |
| TARDBP | 0.37 |
| AZIN1 | 0.37 |
| YARS2 | 0.37 |

| | |
|---|---|
| METTL10 | 0.37 |
| PLEKHA3 | 0.37 |
| TGDS | 0.37 |
| CDC73 | 0.37 |
| ZFR | 0.37 |
| CYCS | 0.37 |
| NASP | 0.37 |
| C14orf126 | 0.37 |
| C4orf43 | 0.37 |
| ENOPH1 | 0.37 |
| SMARCA1 | 0.37 |
| ELAVL1 | 0.37 |
| RBM25 | 0.37 |
| TRMT5 | 0.37 |
| C1orf43 | 0.37 |
| DNAH14 | 0.37 |
| RAD51C | 0.37 |
| BFAR | 0.37 |
| ZNF273 | 0.37 |
| FUNDC2 | 0.37 |
| SLC25A3 | 0.37 |
| CCNB1 | 0.37 |
| CCDC43 | 0.37 |
| L2HGDH | 0.37 |
| PIGF | 0.37 |
| XIAP | 0.37 |
| MRPL13 | 0.37 |
| UBE2O | 0.37 |
| ALKBH1 | 0.37 |
| RNF34 | 0.37 |
| INTS8 | 0.37 |
| IVD | 0.37 |
| RAD54L | 0.37 |
| SSBP1 | 0.37 |
| SUPT7L | 0.37 |
| SKA2 | 0.37 |
| CENPM | 0.37 |
| ABI2 | 0.37 |
| NUDCD2 | 0.37 |
| NUP35 | 0.37 |
| HSP90AB1 | 0.37 |
| ATP13A3 | 0.37 |
| RPS4X | 0.37 |
| CISD2 | 0.37 |
| JOSD1 | 0.37 |
| TBL2 | 0.37 |

| | |
|---|---|
| MTPAP | 0.37 |
| ODF2 | 0.37 |
| DDX54 | 0.37 |
| SDHC | 0.37 |
| NOLC1 | 0.37 |
| MRPL16 | 0.37 |
| DHODH | 0.37 |
| SLC10A3 | 0.37 |
| PIR | 0.36 |
| SNAPC1 | 0.36 |
| FAM161A | 0.36 |
| C1orf107 | 0.36 |
| LCLAT1 | 0.36 |
| NDUFA12 | 0.36 |
| MTX2 | 0.36 |
| FAM127C | 0.36 |
| PPP2R5C | 0.36 |
| NUBPL | 0.36 |
| FAM136A | 0.36 |
| LRRC40 | 0.36 |
| SCML1 | 0.36 |
| PPA1 | 0.36 |
| PAK2 | 0.36 |
| SAAL1 | 0.36 |
| POLE3 | 0.36 |
| OPA1 | 0.36 |
| TTC27 | 0.36 |
| TRMT11 | 0.36 |
| C7orf11 | 0.36 |
| CCT7 | 0.36 |
| IDH3G | 0.36 |
| DNTTIP2 | 0.36 |
| RFC3 | 0.36 |
| TMEM18 | 0.36 |
| PDCD2L | 0.36 |
| FAM60A | 0.36 |
| LSM6 | 0.36 |
| CEP78 | 0.36 |
| RRM1 | 0.36 |
| ZNHIT3 | 0.36 |
| NUSAP1 | 0.36 |
| DUS4L | 0.36 |
| TLK1 | 0.36 |
| URB2 | 0.36 |
| PAPOLG | 0.36 |
| TCERG1 | 0.36 |

| | |
|---|---|
| C14orf142 | 0.36 |
| TSFM | 0.36 |
| CTDSPL2 | 0.36 |
| COMMD2 | 0.36 |
| GPRASP2 | 0.36 |
| PTBP1 | 0.36 |
| SF1 | 0.36 |
| MRPS5 | 0.36 |
| ABCB10 | 0.36 |
| KLHL12 | 0.36 |
| KHSRP | 0.36 |
| POLR2C | 0.36 |
| TH1L | 0.36 |
| UBXN4 | 0.36 |
| FXN | 0.36 |
| ACBD6 | 0.36 |
| YEATS2 | 0.36 |
| PRIM1 | 0.36 |
| EIF2A | 0.36 |
| NVL | 0.36 |
| RPS7 | 0.36 |
| UBE2Q1 | 0.36 |
| MTERF | 0.36 |
| C2orf44 | 0.36 |
| TPI1P2 | 0.36 |
| OXA1L | 0.36 |
| PSMD12 | 0.36 |
| ADORA2B | 0.36 |
| DEK | 0.36 |
| NFU1 | 0.36 |
| RNF41 | 0.36 |
| ZRANB3 | 0.36 |
| KIFC1 | 0.36 |
| RNF2 | 0.36 |
| SKA1 | 0.36 |
| NUDT3 | 0.36 |
| RFWD3 | 0.36 |
| HNRNPAB | 0.36 |
| ABHD3 | 0.36 |
| SLC29A2 | 0.36 |
| UCHL5 | 0.36 |
| TRIM59 | 0.36 |
| MRPL10 | 0.36 |
| NUP88 | 0.36 |
| WDR61 | 0.36 |
| UXS1 | 0.36 |

| | |
|---|---|
| FARSB | 0.36 |
| IFT80 | 0.36 |
| TFRC | 0.36 |
| NAA25 | 0.36 |
| CTAGE5 | 0.36 |
| ELF4 | 0.36 |
| ZNF75D | 0.36 |
| SIRT5 | 0.36 |
| EML4 | 0.36 |
| CBWD2 | 0.36 |
| PDIA6 | 0.36 |
| METTL1 | 0.36 |
| MASTL | 0.36 |
| DCAF7 | 0.36 |
| PLK4 | 0.36 |
| MRPL18 | 0.36 |
| COQ6 | 0.36 |
| SASS6 | 0.36 |
| UBE2C | 0.36 |
| CXorf57 | 0.36 |
| STRBP | 0.36 |
| CENPF | 0.36 |
| TPX2 | 0.36 |
| MTMR1 | 0.36 |
| FAM98B | 0.36 |
| ATF1 | 0.36 |
| PAK1IP1 | 0.36 |
| GTF2A2 | 0.36 |
| TTC8 | 0.36 |
| RABL2B | 0.36 |
| ZNF207 | 0.36 |
| SIVA1 | 0.36 |
| NETO2 | 0.36 |
| MRPS14 | 0.36 |
| SLC25A32 | 0.36 |
| NCAPG2 | 0.36 |
| KIF2C | 0.36 |
| M6PR | 0.36 |
| GIT1 | 0.36 |
| SHCBP1 | 0.36 |
| ATAD2 | 0.36 |
| EWSR1 | 0.36 |
| ACN9 | 0.36 |
| POC1B | 0.36 |
| TMEM216 | 0.35 |
| SUMO1 | 0.35 |

| | |
|---|---|
| ZNF146 | 0.35 |
| PPID | 0.35 |
| C9orf30 | 0.35 |
| HMGXB4 | 0.35 |
| SLC25A15 | 0.35 |
| FAM20B | 0.35 |
| COX16 | 0.35 |
| C1orf96 | 0.35 |
| CWC22 | 0.35 |
| HAUS7 | 0.35 |
| SHMT2 | 0.35 |
| MGC12982 | 0.35 |
| ORC4L | 0.35 |
| ZNF642 | 0.35 |
| PUS7 | 0.35 |
| RRM2 | 0.35 |
| TSPAN6 | 0.35 |
| TSR1 | 0.35 |
| CCDC88A | 0.35 |
| GEMIN4 | 0.35 |
| PFAS | 0.35 |
| SUZ12 | 0.35 |
| TXNL4B | 0.35 |
| ANKMY2 | 0.35 |
| FAM36A | 0.35 |
| LASS2 | 0.35 |
| MARK1 | 0.35 |
| PPIG | 0.35 |
| TADA2A | 0.35 |
| C16orf52 | 0.35 |
| ENY2 | 0.35 |
| WTAP | 0.35 |
| ETV5 | 0.35 |
| GPR89A | 0.35 |
| DNAJC19 | 0.35 |
| C2orf49 | 0.35 |
| WDR89 | 0.35 |
| KIAA0753 | 0.35 |
| LIPT1 | 0.35 |
| SETD3 | 0.35 |
| PABPN1 | 0.35 |
| APOO | 0.35 |
| TTC26 | 0.35 |
| HEATR1 | 0.35 |
| TSR2 | 0.35 |
| PDIA4 | 0.35 |

| | |
|---|---|
| AARS | 0.35 |
| C12orf73 | 0.35 |
| CCT8 | 0.35 |
| C6orf182 | 0.35 |
| NCAPD3 | 0.35 |
| C17orf95 | 0.35 |
| CENPE | 0.35 |
| PIGH | 0.35 |
| NIF3L1 | 0.35 |
| CCDC55 | 0.35 |
| COG2 | 0.35 |
| SNRPE | 0.35 |
| NUP43 | 0.35 |
| NMD3 | 0.35 |
| EXOC6 | 0.35 |
| SLC41A1 | 0.35 |
| FBXO22 | 0.35 |
| CHEK1 | 0.35 |
| EXOSC9 | 0.35 |
| CWF19L1 | 0.35 |
| PANX2 | 0.35 |
| ANAPC5 | 0.35 |
| TAF9B | 0.35 |
| TPI1 | 0.35 |
| ASCC1 | 0.35 |
| VPS29 | 0.35 |
| KIAA0391 | 0.35 |
| SRP54 | 0.35 |
| ESCO2 | 0.35 |
| TAF5L | 0.35 |
| SENP2 | 0.35 |
| PDK1 | 0.35 |
| APOA1BP | 0.35 |
| C7orf44 | 0.35 |
| MUTYH | 0.35 |
| SLC4A5 | 0.35 |
| ZFP1 | 0.35 |
| TWISTNB | 0.35 |
| CKAP2L | 0.35 |
| ELP4 | 0.35 |
| DERA | 0.35 |
| THAP10 | 0.35 |
| PRTFDC1 | 0.35 |
| C14orf138 | 0.35 |
| TRIM37 | 0.35 |
| ATP6AP1 | 0.35 |

| | |
|---|---|
| TIMM9 | 0.35 |
| EIF5 | 0.35 |
| POLR2K | 0.35 |
| ARMCX6 | 0.35 |
| PIGW | 0.35 |
| SAE1 | 0.35 |
| PRPS2 | 0.35 |
| ANAPC10 | 0.35 |
| KCMF1 | 0.35 |
| EME1 | 0.35 |
| ADAM17 | 0.35 |
| ADPGK | 0.35 |
| LTV1 | 0.35 |
| RPL13P5 | 0.35 |
| ATG5 | 0.35 |
| GFM1 | 0.35 |
| TK1 | 0.35 |
| PA2G4 | 0.35 |
| CCT5 | 0.35 |
| CIAO1 | 0.34 |
| EXOSC7 | 0.34 |
| UNG | 0.34 |
| SMC6 | 0.34 |
| PRPF3 | 0.34 |
| PAWR | 0.34 |
| NUP85 | 0.34 |
| E2F4 | 0.34 |
| AKR1E2 | 0.34 |
| KRTCAP3 | 0.34 |
| CDT1 | 0.34 |
| C5orf30 | 0.34 |
| CCDC21 | 0.34 |
| CDCA2 | 0.34 |
| SBDSP1 | 0.34 |
| PDK3 | 0.34 |
| KTN1 | 0.34 |
| PSMC6 | 0.34 |
| CPSF3 | 0.34 |
| TMEM14A | 0.34 |
| MRPL50 | 0.34 |
| RBM12 | 0.34 |
| MSH5 | 0.34 |
| NUDT11 | 0.34 |
| DHX57 | 0.34 |
| PATZ1 | 0.34 |
| CKAP5 | 0.34 |

| | |
|---|---|
| DHX36 | 0.34 |
| FADS1 | 0.34 |
| C14orf145 | 0.34 |
| TTC21B | 0.34 |
| GLOD4 | 0.34 |
| PRMT1 | 0.34 |
| SEMA4F | 0.34 |
| TTLL5 | 0.34 |
| HNRNPA1 | 0.34 |
| SCO1 | 0.34 |
| TIGD1 | 0.34 |
| TAF4 | 0.34 |
| PSME4 | 0.34 |
| KARS | 0.34 |
| CBFB | 0.34 |
| HSP90B1 | 0.34 |
| MEST | 0.34 |
| C6orf72 | 0.34 |
| NCAPG | 0.34 |
| MOSC1 | 0.34 |
| DNAJA3 | 0.34 |
| SF3A3 | 0.34 |
| RPP30 | 0.34 |
| RINT1 | 0.34 |
| ORC3L | 0.34 |
| CDAN1 | 0.34 |
| TGS1 | 0.34 |
| CLK2P | 0.34 |
| MIS12 | 0.34 |
| MAGOH | 0.34 |
| RYK | 0.34 |
| GLRX2 | 0.34 |
| C7orf42 | 0.34 |
| PDCD7 | 0.34 |
| TSNAX | 0.34 |
| LOC728640 | 0.34 |
| EAPP | 0.34 |
| BCORL1 | 0.34 |
| DCTPP1 | 0.34 |
| MLF2 | 0.34 |
| CPSF4 | 0.34 |
| ZNF275 | 0.34 |
| YIPF6 | 0.34 |
| CSPP1 | 0.34 |
| H2AFY2 | 0.34 |
| VEZT | 0.34 |

| | |
|---|---|
| NUFIP1 | 0.34 |
| TARBP1 | 0.34 |
| SLC25A36 | 0.34 |
| DDX10 | 0.34 |
| DGKE | 0.34 |
| PFDN4 | 0.34 |
| RRN3 | 0.34 |
| TAF1B | 0.34 |
| ZNF410 | 0.34 |
| PHKA1 | 0.34 |
| SMN2 | 0.34 |
| CD46 | 0.34 |
| BHLHB9 | 0.34 |
| SNAPIN | 0.34 |
| C14orf135 | 0.34 |
| DENND5A | 0.34 |
| NUP98 | 0.34 |
| ABHD11 | 0.34 |
| ACIN1 | 0.34 |
| RNASEH2A | 0.34 |
| BAZ1B | 0.34 |
| IMMP1L | 0.34 |
| SLMO2 | 0.34 |
| EIF3H | 0.34 |
| MDC1 | 0.34 |
| RPE | 0.34 |
| POLR1B | 0.34 |
| FCF1 | 0.34 |
| LRP8 | 0.34 |
| PSMA1 | 0.34 |
| FAM192A | 0.34 |
| GNPDA1 | 0.34 |
| BCAP31 | 0.34 |
| CS | 0.34 |
| MRPS30 | 0.34 |
| TUBE1 | 0.34 |
| C1D | 0.34 |
| MED9 | 0.34 |
| ZNF326 | 0.34 |
| NDUFAF4 | 0.34 |
| POLR3K | 0.34 |
| TMEM69 | 0.34 |
| SPAST | 0.34 |
| DIAPH3 | 0.34 |
| ORC1L | 0.34 |
| NDUFA1 | 0.34 |

| | |
|---|---|
| PMPCB | 0.34 |
| PPA2 | 0.34 |
| ERGIC2 | 0.34 |
| USP27X | 0.34 |
| AIDA | 0.34 |
| LOC144438 | 0.34 |
| DVL2 | 0.34 |
| DSG2 | 0.34 |
| HS6ST2 | 0.34 |
| LOC646762 | 0.34 |
| RBM12B | 0.34 |
| FAM131A | 0.34 |
| FIP1L1 | 0.34 |
| YLPM1 | 0.34 |
| SART3 | 0.34 |
| RNF13 | 0.34 |
| NARG2 | 0.34 |
| TUBB | 0.34 |
| RSRC1 | 0.34 |
| GPATCH2 | 0.34 |
| GMFB | 0.34 |
| NRF1 | 0.34 |
| RAB15 | 0.34 |
| SENP3 | 0.34 |
| CPSF2 | 0.34 |
| GINS3 | 0.34 |
| YIPF4 | 0.34 |
| TCEAL8 | 0.34 |
| LASS5 | 0.34 |
| HPS3 | 0.34 |
| SMEK1 | 0.34 |
| AATF | 0.34 |
| PGAM5 | 0.34 |
| WBP5 | 0.34 |
| TSEN34 | 0.34 |
| ANGEL2 | 0.34 |
| MYCBP | 0.34 |
| RPL26L1 | 0.34 |
| ECD | 0.34 |
| TKT | 0.34 |
| B4GALNT1 | 0.34 |
| RAD18 | 0.34 |
| MITD1 | 0.34 |
| CCDC77 | 0.34 |
| HSD17B7 | 0.34 |
| ALS2 | 0.34 |

| | |
|---|---|
| TTC19 | 0.34 |
| MPHOSPH10 | 0.34 |
| CCDC7 | 0.34 |
| PPPDE1 | 0.34 |
| LSM14A | 0.34 |
| PFN2 | 0.34 |
| ILF3 | 0.34 |
| CCDC58 | 0.34 |
| FAM49B | 0.34 |
| CENPQ | 0.33 |
| PPIL1 | 0.33 |
| AGPS | 0.33 |
| FKBP4 | 0.33 |
| ETFA | 0.33 |
| DNAJC9 | 0.33 |
| TRAIP | 0.33 |
| ANKRD40 | 0.33 |
| SOCS4 | 0.33 |
| RDH11 | 0.33 |
| CYB5B | 0.33 |
| MBTPS2 | 0.33 |
| UNC50 | 0.33 |
| SFXN4 | 0.33 |
| TBC1D8B | 0.33 |
| ZNF280D | 0.33 |
| FANCD2 | 0.33 |
| SMARCD1 | 0.33 |
| CTPS | 0.33 |
| PPP1R2 | 0.33 |
| CHM | 0.33 |
| FIGNL1 | 0.33 |
| WRN | 0.33 |
| TMEM201 | 0.33 |
| FTSJ2 | 0.33 |
| SLC25A43 | 0.33 |
| ZNHIT6 | 0.33 |
| RHOT1 | 0.33 |
| KNTC1 | 0.33 |
| GTF3C3 | 0.33 |
| PRPF39 | 0.33 |
| MEMO1 | 0.33 |
| RCC1 | 0.33 |
| YWHAG | 0.33 |
| PCBD1 | 0.33 |
| ACSL4 | 0.33 |
| FN3KRP | 0.33 |

| | |
|---|---|
| CXorf40A | 0.33 |
| TBP | 0.33 |
| NIP7 | 0.33 |
| PLDN | 0.33 |
| SNRPD3 | 0.33 |
| B3GALNT2 | 0.33 |
| APITD1 | 0.33 |
| CCAR1 | 0.33 |
| TOMM34 | 0.33 |
| EIF3D | 0.33 |
| TAF9 | 0.33 |
| SDHD | 0.33 |
| MICA | 0.33 |
| MELK | 0.33 |
| C13orf27 | 0.33 |
| MPP6 | 0.33 |
| PIGM | 0.33 |
| RPP40 | 0.33 |
| TMEM209 | 0.33 |
| ABCE1 | 0.33 |
| GART | 0.33 |
| PSMA2 | 0.33 |
| C5orf25 | 0.33 |
| GTPBP4 | 0.33 |
| DCUN1D4 | 0.33 |
| AFMID | 0.33 |
| ZNF77 | 0.33 |
| MARS | 0.33 |
| SFT2D1 | 0.33 |
| ORC5L | 0.33 |
| IFT52 | 0.33 |
| NSMCE2 | 0.33 |
| ANLN | 0.33 |
| ZWINT | 0.33 |
| TMEM5 | 0.33 |
| PRR13 | 0.33 |
| RBM14 | 0.33 |
| LOC646214 | 0.33 |
| TIMELESS | 0.33 |
| SEC62 | 0.33 |
| RNF8 | 0.33 |
| ZDHHC16 | 0.33 |
| GON4L | 0.33 |
| ZNF92 | 0.33 |
| GDAP1 | 0.33 |
| ISG20L2 | 0.33 |

| | |
|---|---|
| TMEM41B | 0.33 |
| C1orf109 | 0.33 |
| MRPL39 | 0.33 |
| AP4S1 | 0.33 |
| CHERP | 0.33 |
| METTL3 | 0.33 |
| ATIC | 0.33 |
| ATP5G2 | 0.33 |
| FBXO11 | 0.33 |
| TPM3 | 0.33 |
| GEMIN6 | 0.33 |
| ZBTB41 | 0.33 |
| ZNF839 | 0.33 |
| BRI3BP | 0.33 |
| C12orf41 | 0.33 |
| TTL | 0.33 |
| TBL1XR1 | 0.33 |
| C6orf167 | 0.33 |
| RPL35A | 0.33 |
| PMS1 | 0.33 |
| EIF3E | 0.33 |
| CDKN2AIPNL | 0.33 |
| SETD4 | 0.33 |
| DDX20 | 0.33 |
| PELP1 | 0.33 |
| MAGEE1 | 0.33 |
| ANGEL1 | 0.33 |
| TCTEX1D2 | 0.33 |
| FAM3A | 0.33 |
| PRMT3 | 0.33 |
| CTPS2 | 0.33 |
| SIX1 | 0.33 |
| FASTKD1 | 0.33 |
| IFT57 | 0.33 |
| POM121 | 0.33 |
| PPPDE2 | 0.33 |
| C2orf64 | 0.33 |
| SP3 | 0.33 |
| AGBL5 | 0.33 |
| NXT2 | 0.33 |
| MCM8 | 0.33 |
| PRDX6 | 0.33 |
| SMPD4 | 0.33 |
| FAM169A | 0.33 |
| EPRS | 0.33 |
| ERCC3 | 0.33 |

| | |
|---|---|
| EBAG9 | 0.33 |
| FLAD1 | 0.33 |
| ZCCHC8 | 0.33 |
| P2RX4 | 0.33 |
| LIG3 | 0.33 |
| FUNDC1 | 0.33 |
| COX15 | 0.33 |
| PDCD5 | 0.33 |
| DGUOK | 0.33 |
| EIF1AX | 0.33 |
| LUC7L2 | 0.33 |
| MKLN1 | 0.33 |
| MAPK1IP1L | 0.33 |
| YBX1 | 0.33 |
| CDC25A | 0.33 |
| TMEM30B | 0.33 |
| ASPH | 0.33 |
| TMEM183A | 0.33 |
| ZNF195 | 0.33 |
| RPL23AP7 | 0.33 |
| SEH1L | 0.33 |
| DUSP11 | 0.33 |
| RCCD1 | 0.33 |
| MCM6 | 0.33 |
| EIF4A2 | 0.33 |
| TMX2 | 0.33 |
| LOC647288 | 0.33 |
| PMS2L1 | 0.33 |
| ELOVL5 | 0.33 |
| HSP90AB2P | 0.33 |
| C14orf19 | 0.33 |
| C12orf65 | 0.33 |
| CORO1C | 0.33 |
| CLN6 | 0.33 |
| SR140 | 0.33 |
| C8orf33 | 0.33 |
| LOC440354 | 0.33 |
| SNRPD1 | 0.33 |
| SIN3A | 0.33 |
| ESD | 0.33 |
| CTCF | 0.33 |
| PSMA4 | 0.33 |
| SEP15 | 0.33 |
| LPCAT3 | 0.33 |
| DIMT1L | 0.32 |
| SFRS6 | 0.32 |

| | |
|---|---|
| ACOT2 | 0.32 |
| DNAJC25 | 0.32 |
| HSP90B3P | 0.32 |
| PHF12 | 0.32 |
| LMNB1 | 0.32 |
| ZNRF3 | 0.32 |
| MTHFD1L | 0.32 |
| ARHGAP11B | 0.32 |
| GMPR2 | 0.32 |
| MRPL33 | 0.32 |
| EFTUD2 | 0.32 |
| LSG1 | 0.32 |
| ARMC8 | 0.32 |
| SMYD5 | 0.32 |
| TMPO | 0.32 |
| KIAA1524 | 0.32 |
| C19orf42 | 0.32 |
| ATP5S | 0.32 |
| NSDHL | 0.32 |
| SKA3 | 0.32 |
| HSPA9 | 0.32 |
| GINS4 | 0.32 |
| CBWD3 | 0.32 |
| FAH | 0.32 |
| RIT1 | 0.32 |
| ZNF680 | 0.32 |
| DIS3L | 0.32 |
| HDGF | 0.32 |
| PITPNB | 0.32 |
| WDSUB1 | 0.32 |
| MESDC2 | 0.32 |
| BIRC5 | 0.32 |
| TCEAL6 | 0.32 |
| COX7B | 0.32 |
| TEX10 | 0.32 |
| SLC19A1 | 0.32 |
| MRPS16 | 0.32 |
| NR2C1 | 0.32 |
| ZNF746 | 0.32 |
| RAD51L1 | 0.32 |
| COIL | 0.32 |
| CPSF7 | 0.32 |
| MYBL2 | 0.32 |
| JKAMP | 0.32 |
| C12orf43 | 0.32 |
| MYO19 | 0.32 |

| | |
|---|---|
| PARL | 0.32 |
| GNL3 | 0.32 |
| ERAL1 | 0.32 |
| ZFP161 | 0.32 |
| MRPL15 | 0.32 |
| C17orf53 | 0.32 |
| ADCY3 | 0.32 |
| RNF7 | 0.32 |
| HAUS4 | 0.32 |
| BCL7A | 0.32 |
| AAAS | 0.32 |
| PPP5C | 0.32 |
| TCEAL3 | 0.32 |
| GEMIN5 | 0.32 |
| KIAA1279 | 0.32 |
| RFWD2 | 0.32 |
| MYNN | 0.32 |
| SUMO3 | 0.32 |
| RUVBL1 | 0.32 |
| ADO | 0.32 |
| C1orf52 | 0.32 |
| SNRPA1 | 0.32 |
| HNRNPF | 0.32 |
| RABEP1 | 0.32 |
| C10orf4 | 0.32 |
| AKT1 | 0.32 |
| SUDS3 | 0.32 |
| MOAP1 | 0.32 |
| RPS6KC1 | 0.32 |
| ANKRD36B | 0.32 |
| SMC3 | 0.32 |
| YWHAQ | 0.32 |
| KIAA1704 | 0.32 |
| MOGS | 0.32 |
| WBP11 | 0.32 |
| ORMDL1 | 0.32 |
| ATXN2 | 0.32 |
| MRPS9 | 0.32 |
| TRUB1 | 0.32 |
| COPS6 | 0.32 |
| EID1 | 0.32 |
| IARS | 0.32 |
| LARP4 | 0.32 |
| TRIM16L | 0.32 |
| PIGC | 0.32 |
| CNPY2 | 0.32 |

| | |
|---|---|
| GPR180 | 0.32 |
| C11orf58 | 0.32 |
| ITGB1BP1 | 0.32 |
| PPM1B | 0.32 |
| KLHDC5 | 0.32 |
| PSMD7 | 0.32 |
| RFXAP | 0.32 |
| PCCB | 0.32 |
| ARSK | 0.32 |
| GSR | 0.32 |
| DPM1 | 0.32 |
| THUMPD3 | 0.32 |
| G6PD | 0.32 |
| WDR44 | 0.32 |
| NPM3 | 0.32 |
| MARS2 | 0.32 |
| LOC401588 | 0.32 |
| RPL23AP82 | 0.32 |
| SAV1 | 0.32 |
| NCEH1 | 0.32 |
| POC5 | 0.32 |
| LOC678655 | 0.32 |
| OSGEPL1 | 0.32 |
| PRPF40A | 0.32 |
| WDR5 | 0.32 |
| TTC9C | 0.32 |
| GTF2H3 | 0.32 |
| IMP4 | 0.32 |
| ALDH18A1 | 0.32 |
| WDR20 | 0.32 |
| PLXNB3 | 0.32 |
| RPL10 | 0.32 |
| GANAB | 0.32 |
| PDSS1 | 0.32 |
| PSIP1 | 0.32 |
| XPOT | 0.32 |
| C12orf45 | 0.32 |
| RNPS1 | 0.32 |
| DNAL1 | 0.32 |
| ZBTB25 | 0.32 |
| TCOF1 | 0.32 |
| SF3B3 | 0.32 |
| RPL36A | 0.32 |
| DHX40 | 0.32 |
| REV1 | 0.32 |
| ARHGAP19 | 0.32 |

| | |
|---|---|
| EXOG | 0.32 |
| RBBP5 | 0.32 |
| ACD | 0.32 |
| NOL8 | 0.32 |
| FOXK2 | 0.32 |
| USP14 | 0.32 |
| C14orf149 | 0.32 |
| FAM120A | 0.32 |
| TMEM106C | 0.32 |
| SOD1 | 0.32 |
| TMEM223 | 0.32 |
| PKP4 | 0.32 |
| PCGF2 | 0.32 |
| C11orf82 | 0.32 |
| MSI2 | 0.32 |
| BAZ1A | 0.32 |
| SLC25A38 | 0.32 |
| DAZAP1 | 0.32 |
| PSMC3IP | 0.32 |
| ANAPC1 | 0.32 |
| IQGAP3 | 0.32 |
| BAT1 | 0.32 |
| THOC4 | 0.32 |
| DCLRE1A | 0.32 |
| TRNT1 | 0.32 |
| RDH14 | 0.32 |
| RABEPK | 0.32 |
| P4HA1 | 0.32 |
| C20orf20 | 0.32 |
| GATC | 0.32 |
| SPC24 | 0.32 |
| COPZ1 | 0.32 |
| DGKG | 0.32 |
| TROVE2 | 0.32 |
| ALOX12P2 | 0.32 |
| UBAP2L | 0.32 |
| GGNBP2 | 0.32 |
| SNRNP40 | 0.32 |
| DBR1 | 0.32 |
| PDCL3 | 0.31 |
| SUZ12P | 0.31 |
| RP9 | 0.31 |
| OSGEP | 0.31 |
| CGRRF1 | 0.31 |
| INTS7 | 0.31 |
| DCAF4 | 0.31 |

| | |
|---|---|
| NAA38 | 0.31 |
| FAIM | 0.31 |
| RPUSD2 | 0.31 |
| ZNF594 | 0.31 |
| KLC2 | 0.31 |
| SEC61A2 | 0.31 |
| OST4 | 0.31 |
| CLK2 | 0.31 |
| PABPC3 | 0.31 |
| F8 | 0.31 |
| HMGB2 | 0.31 |
| FAM120C | 0.31 |
| PDIA5 | 0.31 |
| TMEM67 | 0.31 |
| AKAP1 | 0.31 |
| THNSL1 | 0.31 |
| ASNS | 0.31 |
| METT5D1 | 0.31 |
| PHTF2 | 0.31 |
| CCDC112 | 0.31 |
| SMEK2 | 0.31 |
| PRR3 | 0.31 |
| C6orf162 | 0.31 |
| PPP1R13B | 0.31 |
| FKRP | 0.31 |
| PPP1R2P3 | 0.31 |
| EIF2C3 | 0.31 |
| DERL2 | 0.31 |
| MSTO1 | 0.31 |
| POT1 | 0.31 |
| IGF2BP2 | 0.31 |
| CHST7 | 0.31 |
| ARFGAP3 | 0.31 |
| NACC1 | 0.31 |
| SNRNP200 | 0.31 |
| LRRC57 | 0.31 |
| PHB | 0.31 |
| POLA2 | 0.31 |
| NBN | 0.31 |
| PHKA2 | 0.31 |
| TSSC1 | 0.31 |
| PRPF19 | 0.31 |
| CDC25C | 0.31 |
| C8orf37 | 0.31 |
| TMTC3 | 0.31 |
| SGOL1 | 0.31 |

| | |
|---|---|
| KIAA0406 | 0.31 |
| CABLES2 | 0.31 |
| PIN4 | 0.31 |
| DPH5 | 0.31 |
| HNRNPM | 0.31 |
| RAVER2 | 0.31 |
| NAF1 | 0.31 |
| KIF20A | 0.31 |
| C2orf28 | 0.31 |
| CCDC115 | 0.31 |
| C3orf21 | 0.31 |
| C15orf40 | 0.31 |
| CCDC34 | 0.31 |
| C9orf140 | 0.31 |
| SMS | 0.31 |
| E2F7 | 0.31 |
| FAM3C | 0.31 |
| RAD23A | 0.31 |
| MINPP1 | 0.31 |
| LRP12 | 0.31 |
| AKAP8 | 0.31 |
| RMND1 | 0.31 |
| FANCL | 0.31 |
| EMG1 | 0.31 |
| RBM19 | 0.31 |
| DYNLL2 | 0.31 |
| CDADC1 | 0.31 |
| CMC1 | 0.31 |
| ATXN3 | 0.31 |
| USP45 | 0.31 |
| SCARB1 | 0.31 |
| SFRS8 | 0.31 |
| ITGB3BP | 0.31 |
| MRPL32 | 0.31 |
| ERP29 | 0.31 |
| SF3B1 | 0.31 |
| FBXO4 | 0.31 |
| GNPAT | 0.31 |
| AGMAT | 0.31 |
| CEP57 | 0.31 |
| TRAP1 | 0.31 |
| CLCC1 | 0.31 |
| TAF5 | 0.31 |
| IPO4 | 0.31 |
| RAD1 | 0.31 |
| CEP68 | 0.31 |

| | |
|---|---|
| NAA40 | 0.31 |
| TMEM68 | 0.31 |
| PPIH | 0.31 |
| ZC3HC1 | 0.31 |
| AGPAT5 | 0.31 |
| FTHL3 | 0.31 |
| RPL39L | 0.31 |
| NOP2 | 0.31 |
| SLC1A5 | 0.31 |
| CHCHD7 | 0.31 |
| GATAD1 | 0.31 |
| NANP | 0.31 |
| C21orf45 | 0.31 |
| SMCR7L | 0.31 |
| CAD | 0.31 |
| ANAPC16 | 0.31 |
| MAP1B | 0.31 |
| IFT81 | 0.31 |
| POFUT1 | 0.31 |
| CETN3 | 0.31 |
| SNX6 | 0.31 |
| LOC728758 | 0.31 |
| C1orf112 | 0.31 |
| ZMYM5 | 0.31 |
| MAGEF1 | 0.31 |
| WDR92 | 0.31 |
| SEC22C | 0.31 |
| CABYR | 0.31 |
| TOMM5 | 0.31 |
| TCF20 | 0.31 |
| C2orf60 | 0.31 |
| THOC5 | 0.31 |
| SLC35B2 | 0.31 |
| ECE2 | 0.31 |
| GTPBP3 | 0.31 |
| SSR3 | 0.31 |
| DDX52 | 0.31 |
| TMED2 | 0.31 |
| MTA3 | 0.31 |
| PSAT1 | 0.31 |
| C15orf24 | 0.31 |
| IWS1 | 0.31 |
| PCGF1 | 0.31 |
| PFDN2 | 0.31 |
| PMS2L4 | 0.31 |
| ATAD1 | 0.31 |

| | |
|---|---|
| PPP2R5A | 0.31 |
| DDX39 | 0.31 |
| PPP1R12A | 0.31 |
| PLS3 | 0.31 |
| CNTLN | 0.31 |
| DDX59 | 0.31 |
| BAG5 | 0.31 |
| C1orf57 | 0.31 |
| C2orf47 | 0.31 |
| ZUFSP | 0.31 |
| EXOC5 | 0.31 |
| CENPH | 0.31 |
| CPNE2 | 0.31 |
| PAXIP1 | 0.31 |
| SLBP | 0.31 |
| ERLEC1 | 0.31 |
| THOC3 | 0.31 |
| ACYP1 | 0.31 |
| RPIA | 0.31 |
| HNRPDL | 0.31 |
| SIX4 | 0.31 |
| LMBR1 | 0.31 |
| GEN1 | 0.31 |
| WHSC1 | 0.31 |
| AP3M2 | 0.31 |
| ABCF2 | 0.31 |
| EIF2S1 | 0.31 |
| MTF2 | 0.31 |
| C9orf21 | 0.31 |
| RAB4A | 0.31 |
| TMEM39B | 0.31 |
| FARSA | 0.31 |
| TMEM85 | 0.31 |
| ALKBH5 | 0.31 |
| WDR73 | 0.31 |
| SLC37A3 | 0.31 |
| PRICKLE3 | 0.31 |
| C5orf28 | 0.31 |
| C12orf52 | 0.31 |
| MAPK9 | 0.31 |
| KDM1A | 0.31 |
| VDAC3 | 0.31 |
| EXOSC8 | 0.31 |
| ZC3H18 | 0.31 |
| XPR1 | 0.31 |
| EXOSC1 | 0.31 |

| | |
|---|---|
| NOP56 | 0.31 |
| CDK2AP1 | 0.31 |
| AKR1C3 | 0.31 |
| PDHA1 | 0.31 |
| CSTF1 | 0.31 |
| CEP170 | 0.31 |
| CHAF1A | 0.31 |
| KLRAQ1 | 0.31 |
| PSMD2 | 0.31 |
| ACTR3B | 0.31 |
| C10orf2 | 0.31 |
| ESCO1 | 0.31 |
| MPV17 | 0.31 |
| DYNC2LI1 | 0.31 |
| GAPDH | 0.31 |
| LYRM2 | 0.31 |
| KATNA1 | 0.31 |
| RAG1AP1 | 0.31 |
| TMEM20 | 0.31 |
| RPF2 | 0.31 |
| MLX | 0.31 |
| SMARCD2 | 0.31 |
| RPA1 | 0.31 |
| PIGS | 0.31 |
| TMEM194A | 0.31 |
| PSME3 | 0.31 |
| NAT10 | 0.3 |
| EIF1 | 0.3 |
| FUS | 0.3 |
| POLD2 | 0.3 |
| CENPJ | 0.3 |
| ALS2CR4 | 0.3 |
| PCNP | 0.3 |
| SCLT1 | 0.3 |
| NOL7 | 0.3 |
| ZNF830 | 0.3 |
| RPN2 | 0.3 |
| SAFB | 0.3 |
| KAT2A | 0.3 |
| CNOT10 | 0.3 |
| UBE2NL | 0.3 |
| C15orf57 | 0.3 |
| MEAF6 | 0.3 |
| CHAF1B | 0.3 |
| DSN1 | 0.3 |
| CDC5L | 0.3 |

| | |
|---|---|
| GNL2 | 0.3 |
| TRAPPC6B | 0.3 |
| PIGX | 0.3 |
| POLR2G | 0.3 |
| ITPK1 | 0.3 |
| TOP3A | 0.3 |
| SLC16A14 | 0.3 |
| METTL2B | 0.3 |
| SCARNA12 | 0.3 |
| COQ5 | 0.3 |
| PHOSPHO2 | 0.3 |
| EXOSC3 | 0.3 |
| DMTF1 | 0.3 |
| C6orf57 | 0.3 |
| FAM91A1 | 0.3 |
| GNA13 | 0.3 |
| PIGB | 0.3 |
| C6orf211 | 0.3 |
| HEBP1 | 0.3 |
| BRD7 | 0.3 |
| RBBP8 | 0.3 |
| CCDC91 | 0.3 |
| PSMB4 | 0.3 |
| PDIA3P | 0.3 |
| C16orf87 | 0.3 |
| ANKRD32 | 0.3 |
| NME1 | 0.3 |
| DCK | 0.3 |
| CHID1 | 0.3 |
| ATP5B | 0.3 |
| MCM3 | 0.3 |
| SOX12 | 0.3 |
| PRPF38B | 0.3 |
| PRKCSH | 0.3 |
| ZNF286A | 0.3 |
| HTRA2 | 0.3 |
| ALG10 | 0.3 |
| NUDT5 | 0.3 |
| ATP5C1 | 0.3 |
| UBP1 | 0.3 |
| HMGA1 | 0.3 |
| CASK | 0.3 |
| SNX17 | 0.3 |
| SLC26A10 | 0.3 |
| LMNB2 | 0.3 |
| SAMM50 | 0.3 |

| | |
|---|---|
| MRPS24 | 0.3 |
| PHB2 | 0.3 |
| GPX2 | 0.3 |
| SMURF2 | 0.3 |
| HAUS1 | 0.3 |
| DYNC1LI1 | 0.3 |
| RAB2B | 0.3 |
| GDF11 | 0.3 |
| INTS2 | 0.3 |
| CENPK | 0.3 |
| ATXN2L | 0.3 |
| TM9SF1 | 0.3 |
| CRYZL1 | 0.3 |
| TRAF3 | 0.3 |
| TEX261 | 0.3 |
| PCLO | 0.3 |
| ARIH2 | 0.3 |
| SMARCA5 | 0.3 |
| SRBD1 | 0.3 |
| CDC20 | 0.3 |
| CDC7 | 0.3 |
| OSTC | 0.3 |
| AIFM2 | 0.3 |
| ZNF777 | 0.3 |
| IPO7 | 0.3 |
| QTRTD1 | 0.3 |
| MLH3 | 0.3 |
| PAK1 | 0.3 |
| NSMCE4A | 0.3 |
| SLC33A1 | 0.3 |
| LIN52 | 0.3 |
| MOSPD1 | 0.3 |
| FASTKD2 | 0.3 |
| CDC27 | 0.3 |
| PSMG1 | 0.3 |
| TMEM38B | 0.3 |
| LUC7L3 | 0.3 |
| SNHG1 | 0.3 |
| RBM39 | 0.3 |
| C1orf124 | 0.3 |
| MKRN1 | 0.3 |
| UGGT1 | 0.3 |
| PCMT1 | 0.3 |
| ZBTB26 | 0.3 |
| MED6 | 0.3 |
| POLR3C | 0.3 |

| | |
|---|---|
| RNF220 | 0.3 |
| C19orf48 | 0.3 |
| PRPF18 | 0.3 |
| POLR3H | 0.3 |
| HAUS5 | 0.3 |
| PEX19 | 0.3 |
| FAM55C | 0.3 |
| PMM1 | 0.3 |
| GCLM | 0.3 |
| RPRD1B | 0.3 |
| FBXO7 | 0.3 |
| BTBD1 | 0.3 |
| IMPA1 | 0.3 |
| TFCP2 | 0.3 |
| EHHADH | 0.3 |
| ACTR3 | 0.3 |
| THAP11 | 0.3 |
| PPP1R8 | 0.3 |
| POLA1 | 0.3 |
| UBR5 | 0.3 |
| MPI | 0.3 |
| EIF5A2 | 0.3 |
| NT5DC3 | 0.3 |
| RPL6 | 0.3 |
| SNX27 | 0.3 |
| NCAPD2 | 0.3 |
| RNF219 | 0.3 |
| C9orf80 | 0.3 |
| HIP1 | 0.3 |
| ZNF638 | 0.3 |
| EPS8 | 0.3 |
| C8orf39 | 0.3 |
| RTF1 | 0.3 |
| ZNF7 | 0.3 |
| MRPS28 | 0.3 |
| POC1A | 0.3 |
| ENO2 | 0.3 |
| C7orf28A | 0.3 |
| SCFD1 | 0.3 |
| AKR1B1 | 0.3 |
| HYLS1 | 0.3 |
| BBS5 | 0.3 |
| RIC8B | 0.3 |
| USP36 | 0.3 |
| METTL13 | 0.3 |
| PTGR2 | 0.3 |

| | |
|---|---|
| IDH1 | 0.3 |
| UBTF | 0.3 |
| MTCH2 | 0.3 |
| DCAF17 | 0.3 |
| C14orf147 | 0.3 |
| NAGA | 0.3 |
| TTC32 | 0.3 |
| ATF4 | 0.3 |
| ZNF696 | 0.3 |
| MRPL44 | 0.3 |
| ZNF140 | 0.3 |
| TAF6 | 0.3 |
| ZZZ3 | 0.3 |
| ZXDB | 0.3 |
| KDM3A | 0.3 |
| PFKP | 0.3 |
| HOMEZ | 0.3 |
| XRN2 | 0.3 |
| ATR | 0.3 |
| THAP1 | 0.3 |
| C17orf85 | 0.3 |
| DLST | 0.3 |
| IMPAD1 | 0.3 |
| GCLC | 0.3 |
| ZDHHC17 | 0.3 |
| SNX5 | 0.3 |
| ATP5G3 | 0.3 |
| TUBGCP4 | 0.3 |
| TMEM41A | 0.3 |
| MKS1 | 0.3 |
| MTM1 | 0.3 |
| MRPS7 | 0.3 |
| C3orf17 | 0.3 |
| SETD6 | 0.3 |
| ARMC10 | 0.3 |
| PUS7L | 0.3 |
| ZP3 | 0.3 |
| WDR35 | 0.3 |
| EIF2AK4 | 0.3 |
| FAM83D | 0.3 |
| CAMKK2 | 0.3 |
| ZNF131 | 0.3 |
| ZNF182 | 0.3 |
| GINS1 | 0.3 |
| MUDENG | 0.3 |
| C1orf174 | 0.3 |

| | |
|---|---|
| EP400 | 0.3 |
| CELF1 | 0.3 |
| LACTB2 | 0.3 |
| YWHAH | 0.3 |
| PCBP1 | 0.3 |
| BMS1 | 0.3 |
| RUNDC1 | 0.3 |
| METAP1 | 0.3 |
| SRRM1 | 0.3 |
| RPGRIP1L | 0.3 |
| NECAP1 | 0.3 |
| MTDH | 0.3 |
| GALNT14 | 0.3 |
| SLC39A1 | 0.3 |
| DHX33 | 0.3 |
| MRPS33 | 0.3 |
| PIP5K1A | 0.3 |
| MMACHC | 0.3 |
| CNNM4 | 0.3 |
| MRPS22 | 0.3 |
| DHTKD1 | 0.3 |
| SMYD2 | 0.3 |
| LSM11 | 0.3 |
| SNHG10 | 0.3 |
| GOLIM4 | 0.3 |
| B4GALT3 | 0.3 |
| CLUAP1 | 0.3 |

HNSCC: Head and Neck Squamous Cell Carcinoma, PCC: Pearson correlation coefficient.

### Figures:

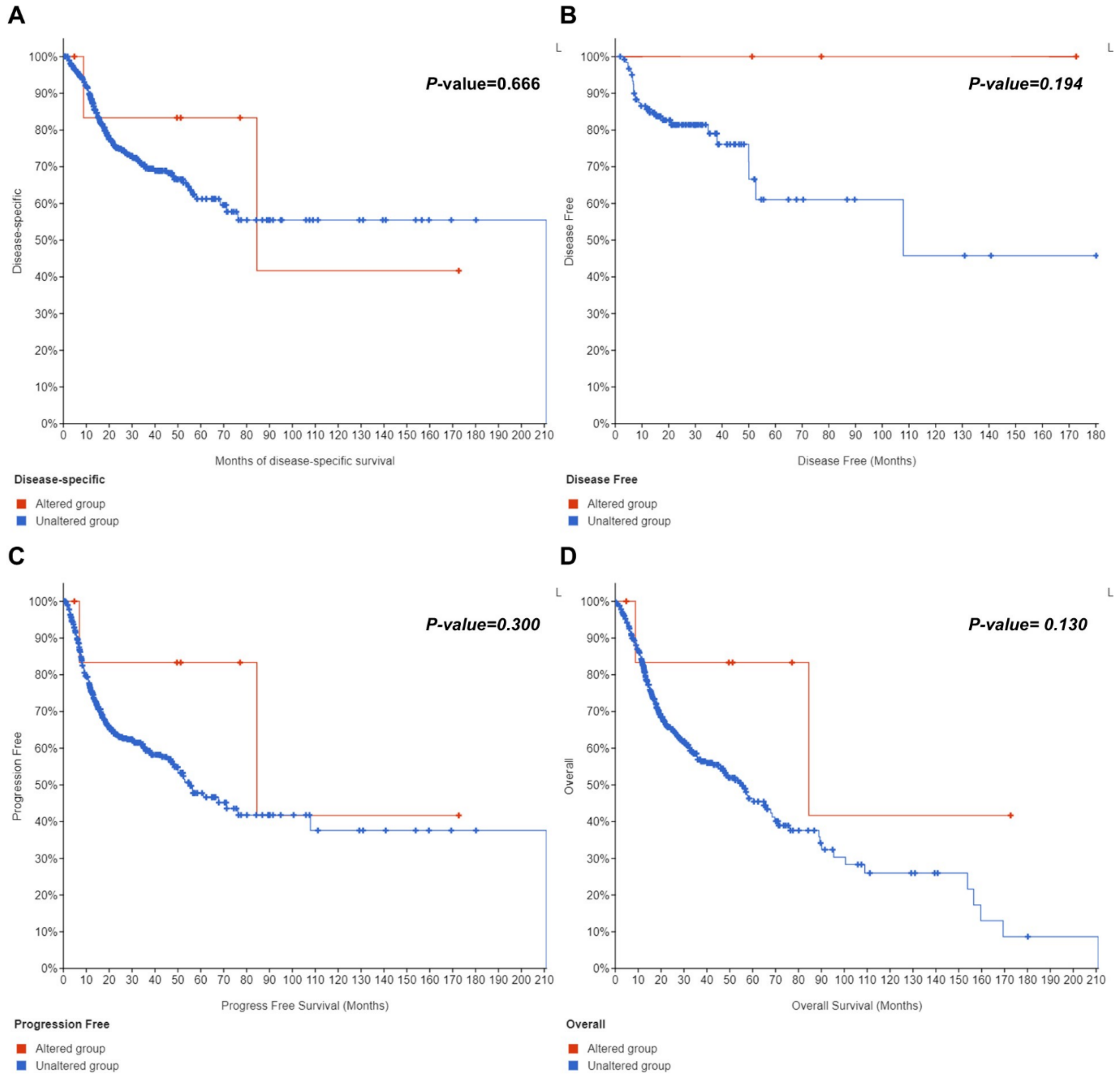


**Figure S1.** The prognostic significance of HPRT1 gene alterations  in HNSCC using the data the cBioPortal database. The association between the genetic alterations of the HPRT1 gene and DSS in patients with HNSCC (A). The relationship between the genetic alterations of the HPRT1 gene and DFS in HNSCC patients (B). The correlation between genetic alterations of the HPRT1 gene and PFS in HNSCC patients (C). The association between the genetic alterations of the HPRT1 gene and OS in patients with HNSCC (D). A log-rank P-value<0.05 was considered as statistically significant. HNSCC: Head and Neck Squamous Cell Carcinoma, DSS: Disease-Specific Survival, DFS: Disease Free Survival, PFS: Progression-Free Survival, OS: Overall Survival.

**Refrences**:

[1] D. Toro-Domínguez, J. Martorell-Marugán, R. López-Domínguez, A. García-Moreno, V. González-Rumayor, M.E. Alarcón-Riquelme, P. Carmona-Sáez, ImaGEO: integrative gene expression meta-analysis from GEO database, Bioinformatics 35(5) (2018) 880-882.
[2] M. Goldman, B. Craft, J. Zhu, D. Haussler, The UCSC Xena system for cancer genomics data visualization and interpretation, AACR, 2017.
[3] P.J. Thul, C. Lindskog, The human protein atlas: A spatial map of the human proteome, Protein Science 27(1) (2018) 233-244.
[4] S.A. Forbes, D. Beare, H. Boutselakis, S. Bamford, N. Bindal, J. Tate, C.G. Cole, S. Ward, E. Dawson, L. Ponting, R. Stefancsik, B. Harsha, C.Y. Kok, M. Jia, H. Jubb, Z. Sondka, S. Thompson, T. De, P.J. Campbell, COSMIC: somatic cancer genetics at high-resolution, Nucleic Acids Research 45(D1) (2016) D777-D783.
[5] E. Cerami, J. Gao, U. Dogrusoz, B.E. Gross, S.O. Sumer, B.A. Aksoy, A. Jacobsen, C.J. Byrne, M.L. Heuer, E. Larsson, Y. Antipin, B. Reva, A.P. Goldberg, C. Sander, N. Schultz, The cBio cancer genomics portal: an open platform for exploring multidimensional cancer genomics data, Cancer Discov 2(5) (2012) 401-4.
[6] B. Gyorffy, A. Lanczky, A.C. Eklund, C. Denkert, J. Budczies, Q. Li, Z. Szallasi, An online survival analysis tool to rapidly assess the effect of 22,277 genes on breast cancer prognosis using microarray data of 1,809 patients, Breast cancer research and treatment 123(3) (2010) 725-31.
[7] B. Gyorffy, P. Surowiak, J. Budczies, A. Lanczky, Online survival analysis software to assess the prognostic value of biomarkers using transcriptomic data in non-small-cell lung cancer, PloS one 8(12) (2013) e82241.
[8] M.V. Kuleshov, M.R. Jones, A.D. Rouillard, N.F. Fernandez, Q. Duan, Z. Wang, S. Koplev, S.L. Jenkins, K.M. Jagodnik, A. Lachmann, Enrichr: a comprehensive gene set enrichment analysis web server 2016 update, Nucleic acids research 44(W1) (2016) W90-W97.
